# Metal Mixtures Mediate the Socioeconomic Gradient in Blood Pressure: A Four-Way Decomposition in a Prospective Rural Bangladeshi Cohort

**DOI:** 10.1101/2025.09.22.25336372

**Authors:** Juwel Rana, Mohammad Hasan Shahriar, Syed Emdadul Haque, Samar Kumar Hore, Tariqul Islam, Golam Sarwar, Muhammad Yunus, Maria Argos, Habibul Ahsan, Jay S. Kaufman

## Abstract

**Background:** The causal mechanisms by which socioeconomic status (SES) affects blood pressure (BP) in low- and middle-income countries (LMICs) remain poorly understood. We examined the effects of SES on BP, and the extent to which disparities in metal mixture exposures mediate these effects among rural Bangladeshi adults.

**Methods:** This study included 5923 participants from the Bangladesh Vitamin E and Selenium Trial (BEST), a prospective cohort followed for six years with repeated BP assessments at baseline and three biennial follow-ups. Baseline exposures included SES indicators: education and agricultural land ownership (socioeconomic position, SEP), and metal mixtures: blood arsenic, lead, selenium, and urinary arsenic. We applied the parametric and mediational g-formula, along with generalized weighted quantile sum regression, to estimate total, direct, and indirect effects of SES on BP outcomes and conduct causal mediation analysis with four-way decomposition.

**Results:** Higher education increased BP, whereas SEP decreased the elevation of BP. Both higher education and SEP lowered metal exposures. Metal mixtures mediated the effects of SES on BP. For example, higher education increased systolic blood pressure (SBP) by 3.53 mmHg (95% CI: 2.23, 4.82), while the pure natural indirect effect showed a protective pathway of –0.44 mmHg (95% CI: –0.62, –0.27) through reduced metals. For SEP, nearly 42% of its protective effect on SBP was mediated by lower metal exposures.

**Conclusions:** Socioeconomic differentials in BP outcomes in rural Bangladesh are partly explained by inequalities in metal mixture exposures. Reducing metal exposures may mitigate SES-related disparities in BP measures in LMICS.

## 1. Introduction

High blood pressure (BP) is the leading modifiable cause of cardiovascular diseases (CVDs) and premature death worldwide, with a disproportionately higher burden in low- and middle-income countries (LMICs), accounting for an estimated 9.15 to 12.1 million deaths in 2022(1,2). In 2020, hypertension affected 31.5% of adults in LMICs (1.04 billion people) compared with 28.5% in high-income countries (349 million people)(3). Similarly, the overall age-standardized prevalence of hypertension in Bangladesh was 26.2% with an increasing trend among adults with high body mass index (BMI) and diabetes(4).

The burden of CVD morbidity and mortality in adults is highly attributable to social and environmental determinants such as education, income, air pollutants, and heavy metal exposures(5–8). Evidence from developed countries suggests that people with lower socioeconomic status (SES), such as education and income, are more vulnerable to high BP and CVD burden due to lack of nutritious food, better housing, healthier lifestyles, and less stressful events(5,9,10). One potential mechanism is through disproportionate exposure to environmental toxins, which elevate BP, dysregulate lipid metabolism, and increase the risk of CVD(11–15). Such unequal exposure patterns reflect broader environmental injustice, where disadvantaged populations carry a disproportionate toxic burden and experience worse health outcomes (5,13,16,17). Consistent with this, several metals, including arsenic, lead, and cadmium, have been positively associated with high BP and CVD outcomes(6,18–22). However, the role of selenium remains inconsistent and requires further prospective studies(20,23). Interestingly, the SES–BP gradient observed in high-income settings appears reversed in LMICs, particularly in South Asia, where higher SES groups often report high BP and prevalence of hypertension(7,17,24–26). While behavioral and lifestyle mechanisms (e.g., diet, obesity, physical activity) are well documented (10,17,25,27–31), metabolic risk factors such as BMI, lipids, glucose, and diabetes have been identified as potential mediators in this pathway(10,24,27,28). However, the contribution of environmental exposures, such as toxic metal mixtures, remains understudied in South Asia, particularly in Bangladesh. Given these contrasting patterns, environmental toxins may play both modifying and mediating roles in the association between SES and BP outcomes. Yet existing evidence on the mediation effect of environmental toxins is largely cross-sectional, and rarely considers environmental mixtures(29,30).

The mediation effect is often examined using traditional methods, assuming no exposure-mediator interaction and no mediator-outcome confounder affected by the exposure(31–33). Recent methodological advancements for causal mediation, such as the mediational g-formula, allow estimation of stochastic analogues of the natural effects under several assumptions. This approach selects mediator values as random draws from the distribution under the alternate exposure group, thereby implementing a stochastic estimator of the natural direct and indirect effects. Thus, it appropriately accounts for exposure-induced mediator–outcome confounding while still allowing for exposure–mediator interaction(34–37). The mediational g-formula was developed initially in a semiparametric form and later extended to a fully parametric framework, which addresses these complexities and offers more robust causal inference(37,38). By applying this counterfactual framework, researchers can better elucidate pathways and mechanisms, thereby informing targeted public health interventions.

Using data from a prospective Bangladeshi cohort, we examined the marginal effects of SES on systolic blood pressure (SBP), diastolic blood pressure (DBP), pulse pressure (PP), and mean arterial pressure (MAP), and assessed the extent to which exposure to a metal mixture mediates socioeconomic differences in BP outcomes. Specifically, we applied VanderWeele’s four-way decomposition method to disentangle the direct, indirect, and interactive effects of SES and metal mixtures on BP(39).

## 2. Methods and Materials

### 2.1 Study population

The study was conducted using data from Bangladesh Vitamin E and Selenium Trial (BEST), a 2×2 factorial, randomized controlled trial that enrolled 7,000 adult permanent residents of arsenic-endemic rural areas (Narayanganj, Comilla, and Noakhali districts under the “Araihazar” study site and Chandpur district under the “Matlab” study site) in Bangladesh. The trial was designed to assess the chemopreventive effects of vitamin E and selenium on arsenical skin cancer. Details of randomization, enrollment procedures, exclusion, and inclusion criteria are described elsewhere(40). Participants aged 25-60 years old were randomized between April 2006 and August 2009 and followed for a 6-year period (i.e., three visits). For this study, the final analytic study population was 5923 individuals who had complete measurements of BP at baseline and during all follow-up visits. We excluded a total of 1077 participants due to taking antihypertensive medication at baseline, lost to follow-up, or death, or without measurements of SBP and DBP. Specific exclusions at each follow-up were 454 participants at follow-up 1 (FU1), 342 at FU2, and 281 at FU3.

### 2.2 Measurements of metal mixture: Mediator

At baseline, blood concentrations of lead (BPb), arsenic (BAs), and selenium (BSe), along with urinary arsenic (UAs), were measured among adults aged 25–60 years. The Generalized Weighted Quantile Sum with Repeated Holdout (gWQSRH) validation index of these four metals was considered as a mediator (M). Urinary arsenic was creatinine-corrected (µg/g Cr). Whole blood concentrations of BPb, BSe, and BAs (µg/L) were measured using an ICP Mass Spectrophotometer (ELAN DRC II, Perkin Elmer), with detection limits of 0.2, 0.4, and 0.2 µg/L, respectively. Non-fasting venous blood samples (10 mL) were collected in vacutainer tubes, stored in 4 °C portable coolers, and processed in a field laboratory within 2–8 hours. UAs (µg/L) was measured using a graphite furnace atomic absorption system (Perkin Elmer; detection limit 2 µg/L). Urinary creatinine was measured using the Jaffe reaction to adjust UAs. Due to skewed distributions, BPb, BSe, BAs, and UAs were log-transformed and used as a mixture in statistical models.

### 2.3 Socioeconomic status (SES) measurements: Exposures

Years of education and ownership of agricultural land (ALO), measured at baseline as SES indicators, were considered as exposures(17,41,42). Years of education were categorized into two groups (0-8 years and 9-17 years of education) based on the country context, allowing for a more meaningful analysis. 8^th^ grade is considered junior school in Bangladesh, and junior school graduates are usually involved in blue-collar jobs. Individuals holding a secondary school certificate (9th-10th grade) or higher education can qualify for white-collar jobs. We consider “0-8 years of education” and “9-17 years of education” at baseline as two hypothetical intervention levels. ALO, one of the main criteria of SEP in rural Bangladesh, was also a binary variable. People are considered wealthy or solvent if they own agricultural lands in rural Bangladesh. Rice is the staple food for Bangladeshi people, and individuals with ALO can cultivate their own food, such as paddy and other grains, in their agricultural lands, which ensures food security and economic solvency. We considered ALO as an indicator of higher SEP and no ALO as lower SEP, representing the second set of hypothetical intervention levels.

### 2.4 BP Measurements: Outcomes

Trained study physicians measured BP at baseline, 2-, 4-, and 6-year follow-up using an automated sphygmomanometer. At each visit, two seated measurements were obtained from the right arm after a 5-minute rest, and their arithmetic mean was used for systolic BP (SBP, mmHg) and diastolic BP (DBP, mmHg). Pulse pressure (PP) was defined as the difference between mean SBP and DBP. Mean arterial pressure (MAP, mmHg) was calculated as DBP + 0.412 × (SBP − DBP), where the factor 0.412 provides greater accuracy in estimating MAP among individuals with uncontrolled hypertension, particularly as PP increases(43). The primary outcomes in this study were SBP, DBP, PP, and MAP measured at the three follow-ups.

### 2.5 Potential Confounders

Baseline covariates were assessed in person using a standardized Bengali questionnaire. These included age (years), gender, betel quid use, smoking status (current, former, never), treatment group (placebo vs. vitamin E & Selenium for skin lesion treatment), occupation (unemployed/retired, farmer/day laborer, homemaker, factory worker, business, other), and antihypertensive medication use. Body mass index (BMI) was calculated from measured height and weight, and diabetes status was based on self-reported physician diagnosis. BMI, diabetes, and medication use were excluded from analyses as potential intermediates in the relationships between SES, metals, and BP(10,27,28). We used a directed acyclic graph (DAG) to identify a minimal set of confounders and intermediate variables in the SES and BP.

### 2.6 Statistical analysis

The study performed univariate and bivariate analyses for each variable. BPb, BSe, BAs, and UAs were log-transformed to minimize the effect of outliers and improve the model assumptions of normal residuals.

#### 2.6.1 Multiple Imputation

At baseline, concentrations of BAs, BSe, and UAs were missing for a small proportion of participants due to insufficient sample volume or machine error (0.5% for BAs and BSe; 0.4% for UAs). BPb was measured in a random subset comprising 14.3% of participants across the four treatment groups, leaving 84.8% missing due to resource constraints. Missingness was assumed to be at random and unrelated to unobserved variables. Multiple imputation by chained equations (MICE) in R was performed, generating 10 imputed datasets with 20 iterations each. The imputation model included outcomes, biomarkers, and all covariates(44)[see Table S1 for details]. Analyses were restricted to participants with measured BP at the 6-year follow-up (N = 5,923). All statistical analyses were conducted across the 10 imputed datasets and pooled following Rubin’s rule(45).

#### 2.6.2 Parametric g-formula

We applied the parametric g-formula in a time-fixed setting to estimate the total effects of education and SEP on BP outcomes at 6 years. Each SES exposure was modeled separately, adjusting for the other SES variable and all confounders. Marginal mean differences in BP outcomes were reported. Indirect effects of SES on BP through the metal mixture were also estimated using the g-formula. Standard errors and 95% confidence intervals (CIs) were obtained via 1,000 bootstrap replications(46).

#### 2.6.3 Weighted Quantile Sum Regression

We estimated the direct effects of SES (A) on BP outcomes (Ys), accounting for the metal mixture (M), using generalized weighted quantile sum regression with repeated holdout (gWQSRH). This approach models the joint effect of correlated environmental chemicals while constraining their effects in the same direction. The gWQSRH method generates a single weighted index of the four metals, standardized into quantiles, representing the overall body burden. Each metal’s relative contribution to the index was estimated through weights reflecting its importance in predicting BP outcomes. We used 100 bootstrap samples with 100 repeated holdout validations to estimate mean differences and metal weights and calculated 95% confidence intervals (CIs) using 1,000 bootstrap replications. gWQSRH regression estimated differences in mean BP per decile increase in the mixture index. Details of the gWQSRH regression are described elsewhere(47,48) and it was implemented using *gWQS* R package (49). The resulting gWQSRH index was then extracted and used as a mediator, which is one of the possible approaches for evaluating the mediation effect of environmental mixtures(30).

#### 2.6.4 Causal Mediation Analysis

Education and SEP were defined as binary exposures (A), the metal mixture index as a continuous mediator (M), and SBP, DBP, PP, and MAP at the third follow-up as continuous outcomes (Y). Age, gender, and treatment group were considered baseline confounders not affected by exposures (V). In contrast, baseline BP, smoking status, betel quid use, and occupation were considered exposure-affected confounders (L) [Figure S2]. Because mediator–outcome confounders affected by exposure may violate standard assumptions, we applied the parametric mediational g-formula, which performs well with continuous mediators, exposure-affected confounders, and high-dimensional settings(37). Under the direct counterfactual framework, we simulated BP outcome at the end of follow-up (6 year) under four joint exposure–mediator distributions and estimated four-way decompositions of the overall effect: controlled direct effect (CDE), randomized analogue of reference interaction (rINTRef), randomized analogue of mediated interaction (rINTMed), randomized analogue of pure natural indirect effect (rPNIE), and total effect (rTE). Monte Carlo simulations and 1,000 bootstrap replications were used to estimate mean differences in BP outcomes and 95% bias-corrected and accelerated confidence intervals. Mediation analysis was implemented using the CMAverse package in R(50).

### 2.7 Secondary Analyses

We repeated the primary analyses for BP outcomes at the 2-year (FU1) and 4-year (FU2) follow-ups to examine temporal patterns of SES and metal mixture effects. In addition, gender-stratified analyses were conducted for BP outcomes over the 6-year follow-up.

### 2.8 Sensitivity Analyses

We assessed robustness by: (i) repeating analyses using complete-case data (excluding imputed BPb values), and (ii) conducting mediation analyses treating individual metals as separate mediators.

## 3. Results of the study

Of the participants, 5,923 (84.6%) completed all four BP measurements through follow-up 3. *Table 1* presents baseline characteristics. The mean age was 41.5 years (SD 10.2), and 62.3% were female. Betel quid use was highly prevalent (56.3%), 18.7% were current smokers, and 60.0% were homemakers. Most participants (83.3%) had ≤8 years of education, and 51.5% owned agricultural land.

**Table 1:**
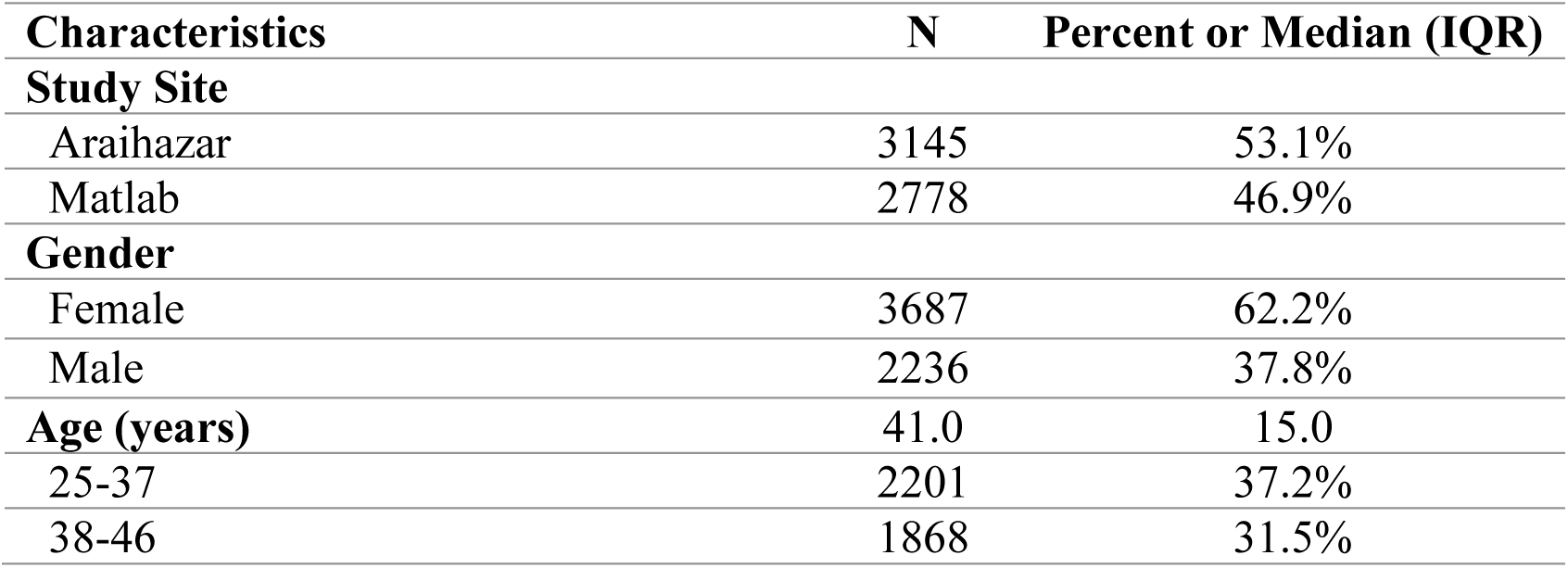
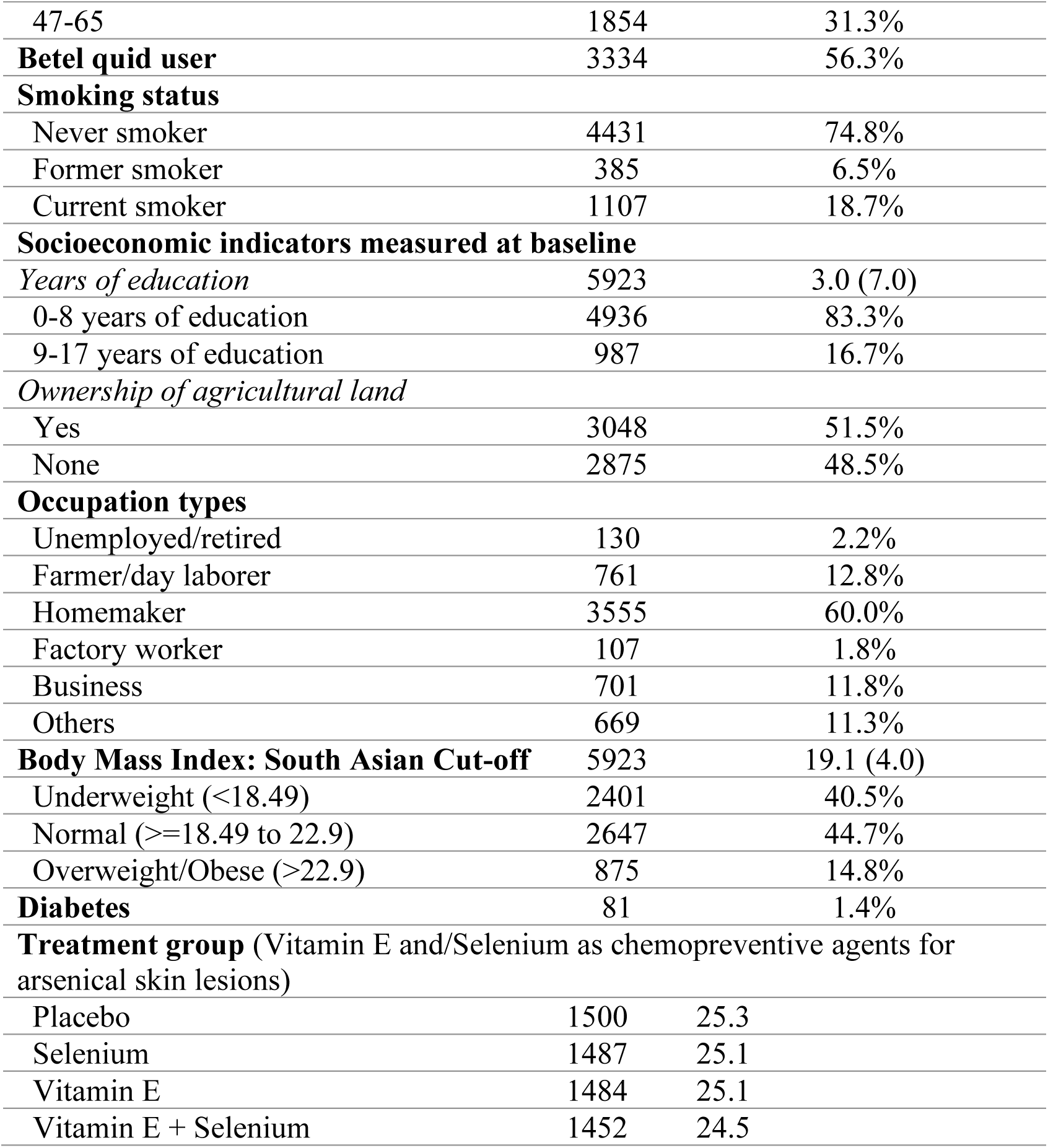
Baseline characteristics of BEST participants (*N*=5923)

*Table 2* shows the distribution of baseline metal concentrations and repeated BP measures over six years of follow-up. Median (25th, 75th percentile) concentrations were 70.3 (35.5, 128.0) µg/L for BPb, 130.0 (115.0, 147.0) µg/L for BSe, 14.8 (4.7, 31.8) µg/L for BAs, and 464.8 (139.6, 1045.0) µg/g Cr for UAs. Distributions were consistent across the complete dataset (compared to the imputed dataset), except for slightly lower values of BPb[Table S2]. SBP, DBP, PP, and MAP increased modestly over a mean follow-up of 6 years [Table 2]. BPb, BSe, BAs, and UAs had weak to strong correlations with a range of [Pairwise Spearman’s rho (r)] -0.02 to 0.94, with the strongest positive correlation between UAs and BAs (r=0.94) and weak positive correlation between BPb and BSe (r=0.18) [Table S3, Fig S1].

**Table 2:** Metal mixture exposure at baseline and BP outcomes at baseline and over six years of follow-up (*N*=5923)

|  | N | Geometric Mean (95% CI) |  |  | Percentile |  |  |
| --- | --- | --- | --- | --- | --- | --- | --- |
|  |  | Gmean | LCI | UCI | 25<sup>th</sup> | Median | 75<sup>th</sup> |
| Baseline Heavy Metals Concentration in Biomarkers |  |  |  |  |  |  |  |
| Imputed Cases Measured at Baseline [pooled estimates across 10 imputed datasets using Rubin's Rules] |  |  |  |  |  |  |  |
| Blood lead (µg/L) | 5923 | 67 | 66 | 69 | 36 | 70 | 128 |
| Blood selenium (µg/L) | 5923 | 130 | 130 | 131 | 115 | 130 | 147 |
| Blood arsenic (µg/L) | 5923 | 12 | 12 | 13 | 5 | 15 | 32 |
| Creatinine-adjusted Urinary arsenic (µg/g Cr) | 5923 | 380 | 368 | 391 | 141 | 468 | 1047 |
| Blood Pressure Outcomes | N | Mean | LCI | UCI | 25<sup>th</sup> | Median | 75<sup>th</sup> |
| Baseline |  |  |  |  |  |  |  |
| SBP (mmHg) | 5923 | 116 | 116 | 117 | 105 | 113 | 123 |
| DBP (mmHg) | 5923 | 75 | 75 | 76 | 70 | 75 | 80 |
| PP (mmHg) | 5923 | 41 | 41 | 41 | 33 | 40 | 46 |
| MAP (mmHg) | 5923 | 92 | 92 | 93 | 82 | 91 | 99 |
| Follow-up 1 |  |  |  |  |  |  |  |
| SBP (mmHg) | 5923 | 113 | 113 | 114 | 100 | 110 | 120 |
| DBP (mmHg) | 5923 | 74 | 74 | 75 | 70 | 76 | 80 |
| PP (mmHg) | 5923 | 39 | 39 | 39 | 30 | 40 | 43 |
| MAP (mmHg) | 5923 | 90 | 90 | 91 | 82 | 89 | 97 |
| Follow-up 2 |  |  |  |  |  |  |  |
| SBP (mmHg) | 5923 | 113 | 113 | 114 | 100 | 110 | 120 |
| DBP (mmHg) | 5923 | 74 | 74 | 75 | 70 | 73 | 80 |
| PP (mmHg) | 5923 | 39 | 39 | 39 | 30 | 40 | 43 |
| MAP (mmHg) | 5923 | 90 | 90 | 91 | 82 | 90 | 97 |
| Follow-up 3 |  |  |  |  |  |  |  |
| SBP (mmHg) | 5923 | 118 | 117 | 118 | 103 | 115 | 130 |
| DBP (mmHg) | 5923 | 77 | 76 | 77 | 70 | 78 | 80 |
| PP (mmHg) | 5923 | 41 | 41 | 42 | 30 | 40 | 50 |
| MAP (mmHg) | 5923 | 94 | 93 | 94 | 84 | 92 | 101 |
| *LCI=Lower confidence interval; UCI=Upper confidence interval. |  |  |  |  |  |  |  |

### 3.1 Total Effects of SES on BP Outcomes and Metal Mixture

We estimated the total marginal effects of education and SEP on BP outcomes after 6 years, and metal mixture at the baseline. As shown in Figure 1, participants with 9–17 years of education had higher SBP, DBP, PP, and MAP after 6 years compared with those with 0–8 years of education. In contrast, participants with higher SEP at baseline had lower SBP, DBP, PP, and MAP after 6 years than those with lower SEP [Figure 1, Table S4].

**Figure 1:**
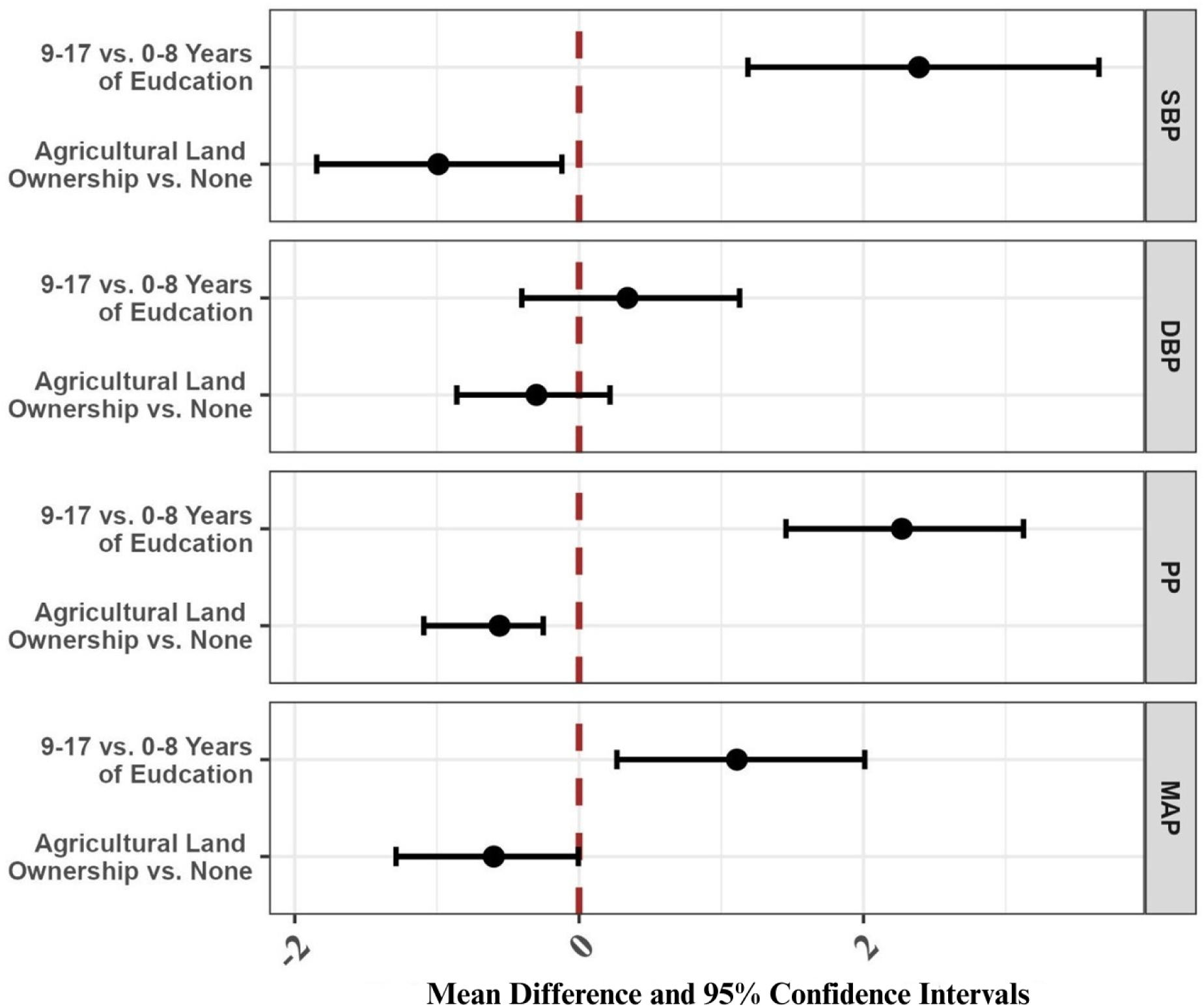
Total marginal effects of socioeconomic indicators on blood pressure outcomes at follow-up 3 (*N*=5923) Note: Mean difference (95% bias-corrected and accelerated bootstrap confidence intervals) in blood pressure (BP) outcomes (mmHg) at follow-up 3 on educational years and ownership of agricultural land (ALO) estimated by parametric g-computation at time-fixed models were adjusted for age, gender, occupation types, baseline BP outcomes, treatment group, betel quid use, and cigarette smoking status as baseline confounders. Education and ALO were also adjusted interchangeably as baseline confounders in their respective models. SBP, systolic blood pressure; DBP, diastolic blood pressure; PP, pulse pressure; MAP, mean arterial pressure.

Higher education and SEP were associated with lower metal mixture concentration (mediator: WQS Index of four metals in deciles) of –0.57 (95% CI: –0.72, –0.43) and –0.28 (95% CI: –0.38, –0.17) deciles, respectively. In other words, higher SES was associated with lower metal mixture exposure [Table S6].

### 3.2 Direct Effects of SES and Metal Mixture on BP Outcomes

*Figures 2*-*3* show the effects of SES and WQS metal mixture Index on BP outcomes at the third follow-up, as well as the contribution of each metal. Participants with 9–17 years of education consistently had higher BP at 6 years, even after including the mediator [BPb, BSe, BAs, and UAs] in the gWQSRH models. In contrast, higher SEP was consistently associated with lower BP after 6 years. The WQS metal mixture index was positively associated with BP outcomes, with each decile increase corresponding to higher BP(SBP_MD_: 0.63; 95% CI: 0.42, 0.85; DBP_MD_: 0.63; 95% CI: 0.51, 0.75; PP_MD_: 0.23; 95% CI: 0.10, 0.36; MAP_MD_: 0.62; 95% CI: 0.47, 0.76). The estimated weights from the gWQSRH reveal that BPb and BSe contributed most to the metal mixture Index (94% to 99%) [Fig 3].

**Figure 2:**
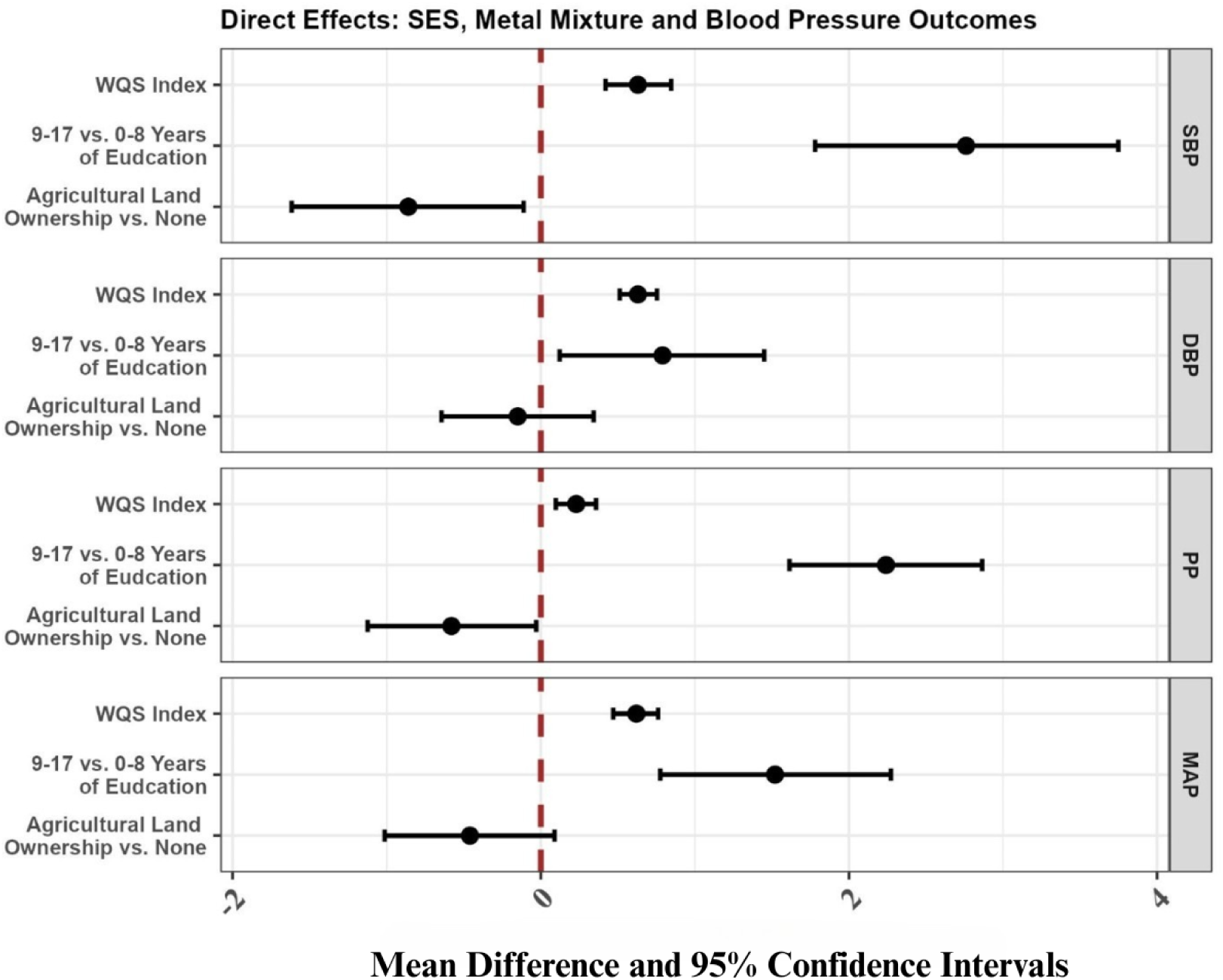
Direct effects of socioeconomic indicators, and metal mixture exposure at baseline on blood pressure outcomes at follow-up 3 (*N*=5923) Note: Mean difference and 95% bootstrap confidence intervals in blood pressure outcomes (mmHg) at follow-up 3 per decile increase in the gWQSRH metal mixture index and on educational years and ownership of agricultural land estimated by the Repeated Holdout Validation (100) WQS regression models were adjusted for age, gender, occupation types, baseline BP, treatment group, betel quid use, and cigarette smoking status. Estimates were pooled across 10 multiple imputed datasets using Rubin’s Rules. SBP, systolic blood pressure; DBP, diastolic blood pressure; PP, pulse pressure; MAP, mean arterial pressure.

**Figure 3:**
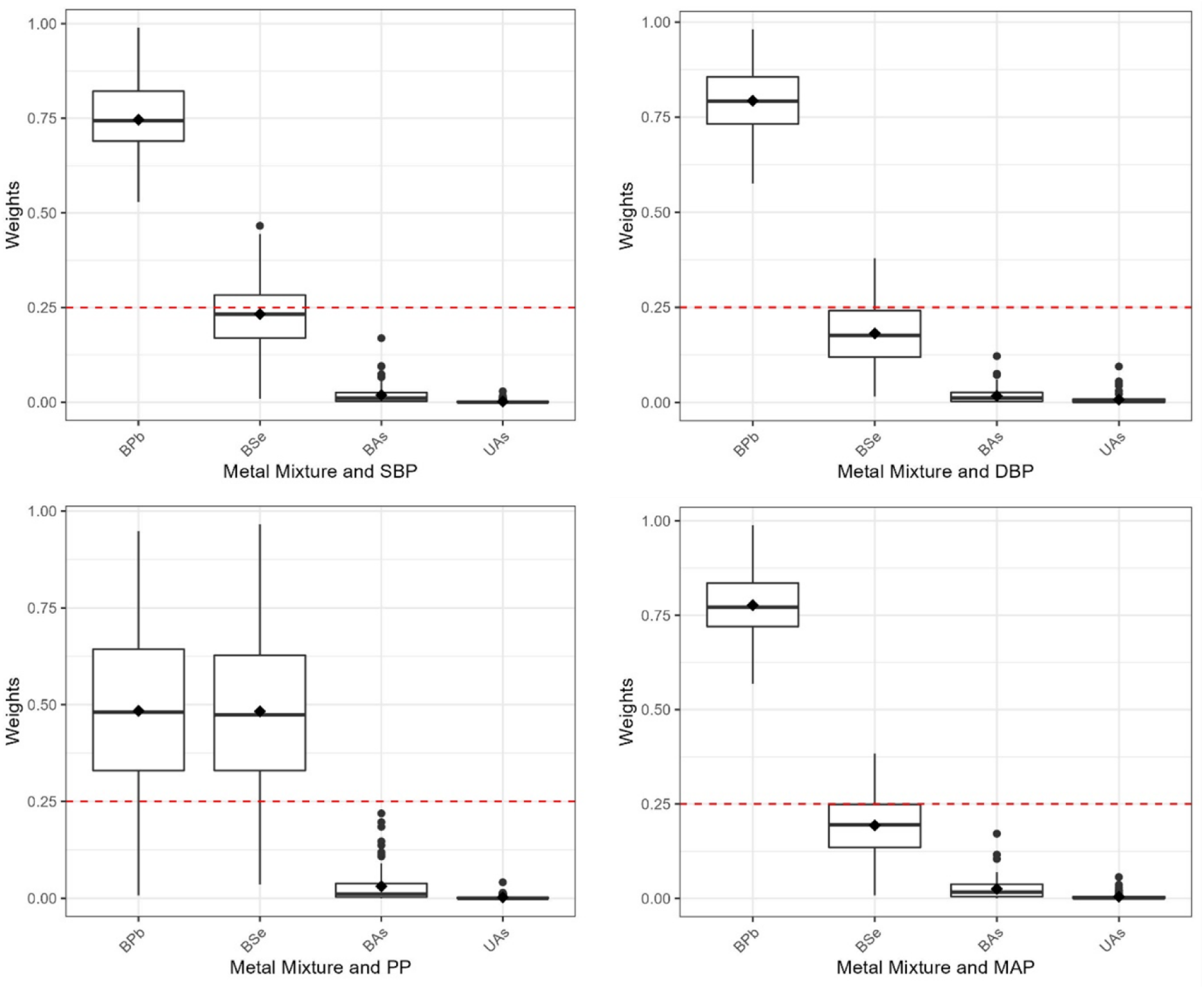
Distribution of each heavy metal weight in Repeated Holdout WQS Index for blood pressure outcomes (*N*=5923) Note: The dashed red line represents the prespecified cutoff to discriminate between significant and non-significant weights equal to the inverse number of elements in the mixture. Repeated Holdout Validation (100) Weighted Quantile Sum (WQS) regression models with socioeconomic indicators, and metal mixture exposure at baseline were adjusted for age, gender, occupation types, baseline BP, treatment group, betel quid use, and cigarette smoking status.

### 3.3 Mediation Effect of Metal Mixture: Four-Way Decomposition

*Table 3* presents the four-way decomposition of the causal mediation effect of the baseline metal mixture, considering mediator-outcome confounders affected by exposure to SBP, DBP, PP, and MAP at follow-up 3, as estimated using the mediational g-formula. The rTE indicates that 9-17 years of education at baseline increased 3.53 mmHg (95% CI: 2.23, 4.82) SBP after six years of follow-up. The CDE, fixing mediator metal mixture (M)=m/1^st^ deciles, was 2.96 mmHg SBP (95% CI: 0.49, 5.55), suggesting that 84% of the effect remained after accounting for the mediator. The rINTRef was 1.15 mmHg (95% CI: -0.97, 3.30), and rINTMed was -0.14 mmHg (95% CI: -0.42, 0.11), indicating that the education–metal mixture interaction effect appeared more important than the mediated interaction, although both were null. The rPNIE reveals that education can reduce - 0.44 mmHg (95% CI: -0.62, -0.27) SBP at six years of intervening on metal mixture only. The rPNIE showed that education reduced SBP by –0.44 mmHg (95% CI: –0.62, –0.27) through the pathway of metal mixture alone. The opposite directions of the TE/rTNDE and rPNIE suggest that the seemingly protective mediated effect of education via metal mixture may partially mask its detrimental total and direct effects on SBP. Similar patterns were observed for DBP, PP, and MAP.

**Table 3:**
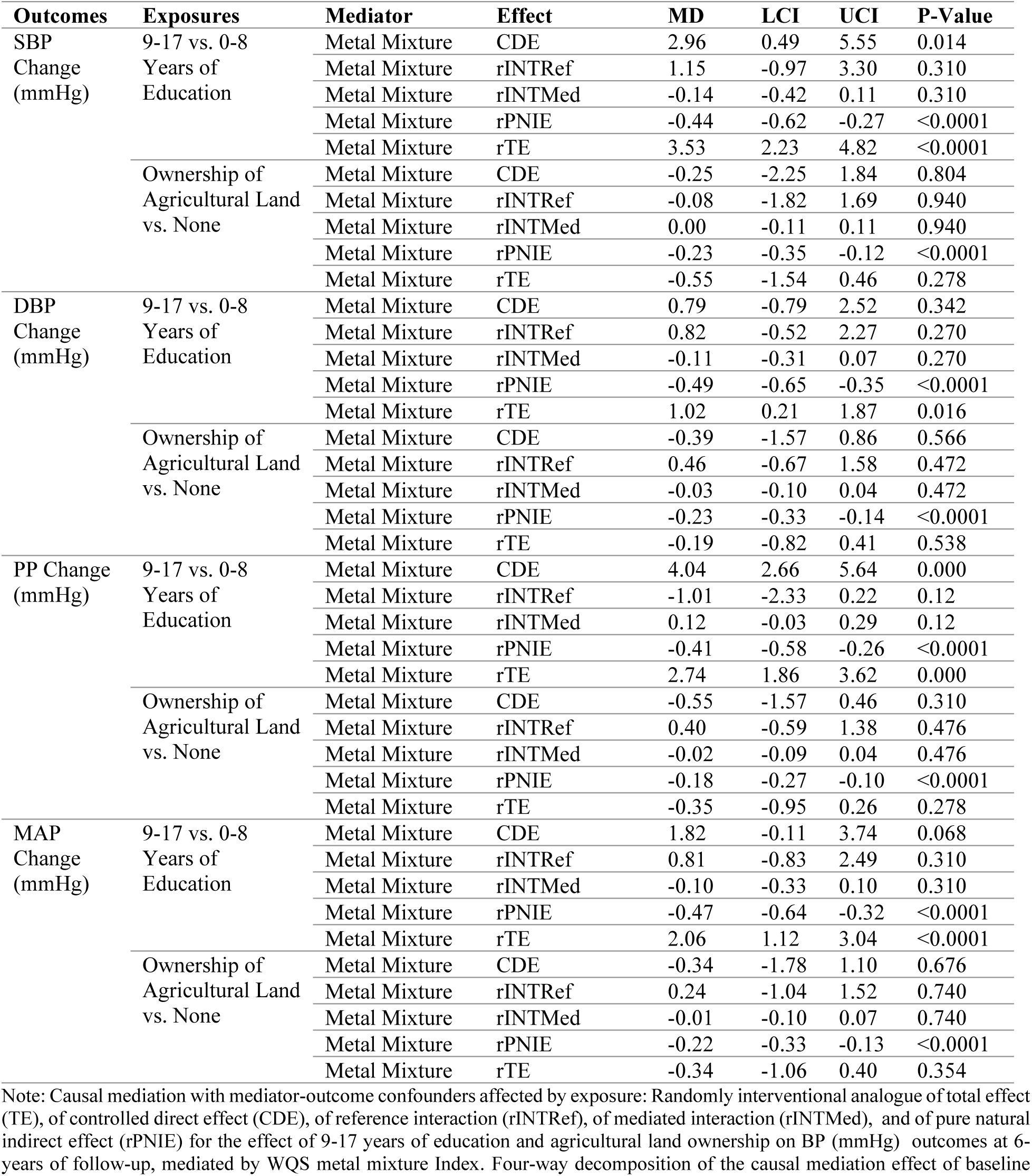
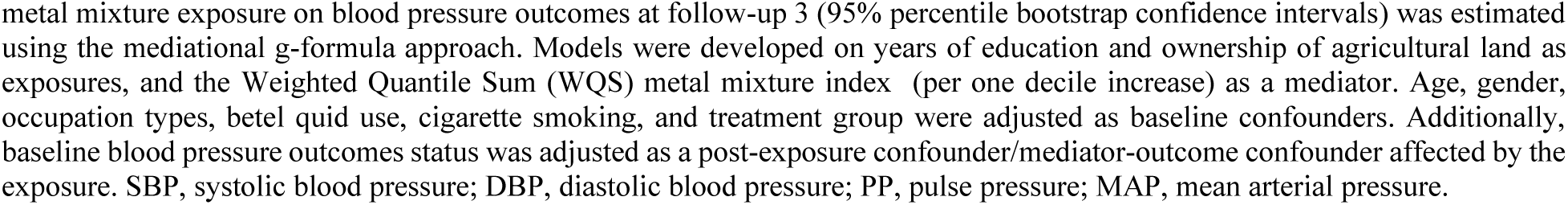
Randomly interventional analogue of TE, of CDE, of rINTRef, of rINTMed, and of rPNIE for the effect of education and ALO for 6 years on BP outcomes, mediated by WQS metal mixture Index (*N*=5923).

| Outcomes | Exposures | Mediator | Effect | MD | LCI | UCI | P-Value |
| --- | --- | --- | --- | --- | --- | --- | --- |
| SBP Change (mmHg) | 9-17 vs. 0-8 Years of Education | Metal Mixture | CDE | 2.96 | 0.49 | 5.55 | 0.014 |
|  |  | Metal Mixture | rINTRef | 1.15 | -0.97 | 3.30 | 0.310 |
|  |  | Metal Mixture | rINTMed | -0.14 | -0.42 | 0.11 | 0.310 |
|  |  | Metal Mixture | rPNIE | -0.44 | -0.62 | -0.27 | <0.0001 |
|  |  | Metal Mixture | rTE | 3.53 | 2.23 | 4.82 | <0.0001 |
|  | Ownership of Agricultural Land vs. None | Metal Mixture | CDE | -0.25 | -2.25 | 1.84 | 0.804 |
|  |  | Metal Mixture | rINTRef | -0.08 | -1.82 | 1.69 | 0.940 |
|  |  | Metal Mixture | rINTMed | 0.00 | -0.11 | 0.11 | 0.940 |
|  |  | Metal Mixture | rPNIE | -0.23 | -0.35 | -0.12 | <0.0001 |
|  |  | Metal Mixture | rTE | -0.55 | -1.54 | 0.46 | 0.278 |
| DBP Change (mmHg) | 9-17 vs. 0-8 Years of Education | Metal Mixture | CDE | 0.79 | -0.79 | 2.52 | 0.342 |
|  |  | Metal Mixture | rINTRef | 0.82 | -0.52 | 2.27 | 0.270 |
|  |  | Metal Mixture | rINTMed | -0.11 | -0.31 | 0.07 | 0.270 |
|  |  | Metal Mixture | rPNIE | -0.49 | -0.65 | -0.35 | <0.0001 |
|  |  | Metal Mixture | rTE | 1.02 | 0.21 | 1.87 | 0.016 |
|  | Ownership of Agricultural Land vs. None | Metal Mixture | CDE | -0.39 | -1.57 | 0.86 | 0.566 |
|  |  | Metal Mixture | rINTRef | 0.46 | -0.67 | 1.58 | 0.472 |
|  |  | Metal Mixture | rINTMed | -0.03 | -0.10 | 0.04 | 0.472 |
|  |  | Metal Mixture | rPNIE | -0.23 | -0.33 | -0.14 | <0.0001 |
|  |  | Metal Mixture | rTE | -0.19 | -0.82 | 0.41 | 0.538 |
| PP Change (mmHg) | 9-17 vs. 0-8 Years of Education | Metal Mixture | CDE | 4.04 | 2.66 | 5.64 | 0.000 |
|  |  | Metal Mixture | rINTRef | -1.01 | -2.33 | 0.22 | 0.12 |
|  |  | Metal Mixture | rINTMed | 0.12 | -0.03 | 0.29 | 0.12 |
|  |  | Metal Mixture | rPNIE | -0.41 | -0.58 | -0.26 | <0.0001 |
|  |  | Metal Mixture | rTE | 2.74 | 1.86 | 3.62 | 0.000 |
|  | Ownership of Agricultural Land vs. None | Metal Mixture | CDE | -0.55 | -1.57 | 0.46 | 0.310 |
|  |  | Metal Mixture | rINTRef | 0.40 | -0.59 | 1.38 | 0.476 |
|  |  | Metal Mixture | rINTMed | -0.02 | -0.09 | 0.04 | 0.476 |
|  |  | Metal Mixture | rPNIE | -0.18 | -0.27 | -0.10 | <0.0001 |
|  |  | Metal Mixture | rTE | -0.35 | -0.95 | 0.26 | 0.278 |
| MAP Change (mmHg) | 9-17 vs. 0-8 Years of Education | Metal Mixture | CDE | 1.82 | -0.11 | 3.74 | 0.068 |
|  |  | Metal Mixture | rINTRef | 0.81 | -0.83 | 2.49 | 0.310 |
|  |  | Metal Mixture | rINTMed | -0.10 | -0.33 | 0.10 | 0.310 |
|  |  | Metal Mixture | rPNIE | -0.47 | -0.64 | -0.32 | <0.0001 |
|  |  | Metal Mixture | rTE | 2.06 | 1.12 | 3.04 | <0.0001 |
|  | Ownership of Agricultural Land vs. None | Metal Mixture | CDE | -0.34 | -1.78 | 1.10 | 0.676 |
|  |  | Metal Mixture | rINTRef | 0.24 | -1.04 | 1.52 | 0.740 |
|  |  | Metal Mixture | rINTMed | -0.01 | -0.10 | 0.07 | 0.740 |
|  |  | Metal Mixture | rPNIE | -0.22 | -0.33 | -0.13 | <0.0001 |
|  |  | Metal Mixture | rTE | -0.34 | -1.06 | 0.40 | 0.354 |

Note: Causal mediation with mediator-outcome confounders affected by exposure: Randomly interventional analogue of total effect (TE), of controlled direct effect (CDE), of reference interaction (rINTRef), of mediated interaction (rINTMed), and of pure natural indirect effect (rPNIE) for the effect of 9-17 years of education and agricultural land ownership on BP (mmHg) outcomes at 6-years of follow-up, mediated by WQS metal mixture Index. Four-way decomposition of the causal mediation effect of baseline
metal mixture exposure on blood pressure outcomes at follow-up 3 (95% percentile bootstrap confidence intervals) was estimated using the mediational g-formula approach. Models were developed on years of education and ownership of agricultural land as exposures, and the Weighted Quantile Sum (WQS) metal mixture index (per one decile increase) as a mediator. Age, gender, occupation types, betel quid use, cigarette smoking, and treatment group were adjusted as baseline confounders. Additionally, baseline blood pressure outcomes status was adjusted as a post-exposure confounder/mediator-outcome confounder affected by the exposure. SBP, systolic blood pressure; DBP, diastolic blood pressure; PP, pulse pressure; MAP, mean arterial pressure.

In contrast, the rTE of higher SEP indicated a reduction of –0.55 mmHg (95% CI: –1.54, 0.46) in SBP, which attenuated to null after accounting for the education–metal mixture interaction and the baseline SBP confounder affected by the exposures. The rINTRef and rINTMed were negligible and imprecise. However, the rPNIE showed that higher SEP reduced SBP by –0.23 mmHg (95% CI: –0.35, –0.12) through the pathway of metal mixture, accounting for 41% of the TE. The directions of rTE/rTNDE and rPNIE were consistent. Similar effects of SEP on DBP, PP, and MAP were observed through mediation by the metal mixture.

### 3.4 Secondary Analyses

Results from the 2- and 4-year follow-ups showed consistent longitudinal increases in the effects of SES indicators and the metal mixture mediator on BP outcomes, supporting the main findings [Tables S4–S7a]. Specifically, rPNIE, rINTRef, and TE increased over the six years of follow-up, confirming the long-term influence of SES and the metal mixture on BP outcomes [Table S7a]. Gender-stratified analyses indicated that the effects of education on BP outcomes were greater among males, while the effects of SEP were smaller compared to females [Table S9a]. Conversely, the rPNIE of education on BP outcomes was higher among females than males, suggesting that the effects of SES and metal mixture may differ by gender.

### 3.5 Sensitivity Analyses

Concentrations of individual metals and their correlations in complete cases were similar to the imputed dataset, though BPb levels were slightly lower in the imputed data [Tables S1–S2]. Complete-case analyses (n = 900) of the four metals (BPb, BSe, BAs, UAs) produced results consistent with multiple imputation models, although most estimates were higher [Tables S4–S6, S8a, S10a]. Gender- and age-adjusted effects of individual metals varied substantially, and standard errors were inflated in models with mutual adjustment for all four metals, confirming co-confounding and multicollinearity, and supporting the use of a mixture approach. When metals were modeled as multiple mediators, the mediational g-formula failed to handle their high correlation, distorting estimates, further justifying the use of the mixture index. Quadratic terms in WQS models showed no evidence of non-linearity. BMI was not adjusted for, as it is an intermediate in the SES–BP and metals–BP pathways. Adjustment for study site and BP-lowering medication slightly reduced effect estimates. Additional analyses accounting for baseline BP, occupation, betel quid use, and smoking status as post-exposure confounders/mediator–outcome confounders, using both imputed and complete data, yielded similar findings: rTE and CDE for higher education and SEP increased, while rPNIE for the metal mixture was slightly attenuated [Tables 7b–10b]. Overall, results from imputed data were conservative compared with complete-case analyses, but conclusions remained unchanged, robust, and strengthened by sensitivity analyses.

## 4. Discussion

Our study demonstrates that higher education increased SBP, DBP, PP, and MAP over six years of follow-up among rural Bangladeshi adults. The controlled direct effect indicated that a substantial proportion of the total effect of education on BP outcomes (up to 89%) remained even when fixing the metal mixture at the first decile. The indirect effect of education through metals was protective: higher education lowered metal exposures, which in turn slightly reduced average BP. Although the education–metal mixture interaction effect exceeded the mediation effect, interaction term estimates were imprecise. By contrast, higher SEP, measured by ALO, reduced the risk of elevated BP. Most of the protective total effect of SEP (up to 82%) on SBP, DBP, PP, and MAP was explained by the indirect pathway, suggesting that higher SEP reduces the risk of elevated BP primarily by reducing inequalities in metal exposures. The controlled direct effect of SEP was small and imprecise, while the total protective effect appeared driven mainly by the mediation and interaction of metals, especially lead and selenium.

The relationship between higher education and BP outcomes revealed slight gender differences, with the relationship between education and metal mixtures being stronger among males than females. In contrast, both the total effect and pure natural indirect effect of SEP were higher among females(5,51). Overall, education and SEP exerted effects on BP outcomes that differed in both magnitude and direction. The distinct mechanisms observed further justify our treatment of education and SEP as separate SES indicators(41,52).

We found that higher education elevated BP outcomes in LMICs, with stronger effects among males in South Asia, including Bangladesh, which are consistent with previous studies(5,7,17,24,42,53). This pattern contrasts with findings from high-income countries, where higher education is generally protective(11,41,54). The total and direct effects of higher education were detrimental, increasing SBP, DBP, PP, and MAP over six years of follow-up. In contrast, the indirect effect mediated through the metal mixture was protective, reflecting lower exposures among more educated adults. This opposing directionality suggests that the protective pathway of reduced metal exposure partially offset, but did not eliminate, the overall harmful influence of higher education on BP. Lifestyle factors linked to higher education in LMICs, such as reduced physical activity, greater consumption of calorie-dense diets, high salt intake, processed foods, and stress, may drive the detrimental pathway(17,25,53,55). Gender differences may further reflect distinct lifestyle behaviors and social roles between men and women in rural South Asian settings(7,24,56,57). At the same time, occupational and environmental advantages of education reduce metal exposures and modestly counterbalance these harmful effects. Together, this decomposition helps explain why higher education appears to elevate BP in South Asia, in contrast to the protective associations observed in high-income settings.

Education had both interacting and mediating effects with the metal mixture on BP outcomes, consistent with prior studies(11,13). Evidence from the USA shows that individuals with higher education tend to have lower metal mixture concentrations than those with less education (5,29,58). In our study, the mixture, particularly lead, and selenium, was associated with increased SBP, DBP, PP, and MAP, aligning with previous mixture and single-pollutant findings(19,59–62). Proposed mechanisms include acute autonomic responses and stimulation of oxidative stress and inflammation, leading to increased reactive oxygen species (ROS) and reduced antioxidant defenses (14,22,63,64). These pathways may elevate BP and CVD risk by disrupting ROS balance, promoting weight gain, and altering lipid metabolism(59,65). Recent meta-analyses also reported that heavy metals, including arsenic and lead, significantly increase CVD risk through higher SBP and renal dysfunction(6,66,67). Our findings suggest that the effects of education on BP outcomes are partly explained by metal mixture exposures, which contribute to socioeconomic disparities in BP-related morbidity and mortality.

In contrast to education, higher SEP was protective against elevated BP outcomes, although most estimates were imprecise except for the indirect pathway. The strong indirect effect indicates that the protective effect of SEP was fully mediated by the metal mixture, as those with lower SEP in rural areas are more likely to be exposed to metals and thus have higher BP. Rural adults with lower SEP often rely on agricultural work, either cultivating their own land or working as laborers. Soil and air are key sources of Pb exposure through pesticide use and contamination from industrial activities such as lead–acid batteries, paint, and ceramics(68–70). Additional dietary sources of Pb include rice and widely consumed spices such as turmeric and red chili powder in rural Bangladesh(59,71,72).

The effects of SEP and education on SBP, DBP, PP, and MAP in this study were explained by both mediation and interaction through the metal mixture. Higher SEP and education were associated with lower risks of metal mixture exposure, consistent with previous studies in the US showing that individuals with higher SES had lower urinary and blood metal concentrations(11). However, one US study reported that the poverty–income ratio (PIR) was positively associated with arsenic but inversely associated with lead and selenium(58). Our findings, consistent with prior studies in the US(11,13,58,73), highlight socioeconomic inequalities in metal mixture exposures and raise concerns about environmental justice in Bangladesh. Individuals with higher education and SEP are more likely to avoid metal exposures, such as from agricultural activities. In comparison, those with lower education and SEP face higher risks through agricultural labor, battery recycling, garment work, and other industries(68,72,74). Higher SEP may also reduce household exposures, as families often employ domestic workers for household chores in rural Bangladesh. At the same time, unequal access to safe drinking water further exacerbates environmental injustices in arsenic-endemic areas(20). Our findings demonstrate that the protective role of SEP on BP outcomes was fully mediated by the metal mixture, suggesting that interventions to reduce metal exposures rather than changes in SEP, per se, could yield substantial benefits.

Our four-way decomposition relied on several strong causal assumptions, including conditional exchangeability, positivity, and SUTVA. For the stochastic CDE, we assumed no unmeasured confounding of SES–BP, SES–metal exposures, or metals–BP associations. To address mediator– outcome confounders affected by exposure, we used randomized analogues of effects based on stochastic interventions. The mediated and reference interaction components further required strong cross-world counterfactual assumptions, comparing BP outcomes under hypothetical scenarios where individuals retain their observed metal exposures while experiencing counterfactual socioeconomic conditions(37–39).

### 4.1 Strengths and Limitations

A key strength of this study is the use of the parametric mediational g-formula, which accounts for exposure–mediator and mediator–outcome confounders affected by the exposure, as well as exposure–mediator interaction. This counterfactual framework allowed four-way decomposition of the total effect of education and SEP on BP outcomes, as well as partitioning of effects attributable to mediation by the metal mixture. Importantly, ignoring post-exposure confounding produced a significant total effect of SEP on BP (Figure 1), which was diminished in magnitude after proper adjustment (Table 3). Another strength is the use of gWQSRH to model metals as a mixture, avoiding multicollinearity, co-confounding, and unstable estimates. Sensitivity analyses reinforced the validity of treating metals as a mixture rather than multiple mediators. This large, prospective cohort from Bangladesh, combined with a robust analytic approach, clarifies paradoxical associations, revealing that education is not uniformly harmful but exerts both protective and adverse influences. Such insights are essential for designing interventions that reduce socioeconomic inequalities in environmental mixtures and preserve the environmental benefits of education while addressing behavioral risks that contribute to elevated BP in LMICs. This study has several limitations. First, SES was measured with two binary indicators, which may not fully capture gradients, may interact, and are subject to misclassification. Indeed, 83% of the adult population was categorized as having low education, leaving substantial heterogeneity in that category. We also lacked information on land size, which limited the precision of the SEP. As a result, SES effects on BP may be underestimated, though sensitivity analyses using education as a continuous variable gave similar results. Second, BPb, BSe, BAs, and UAs were measured only at baseline, preventing assessment of time-varying effects. Metals were measured in blood, which is not the gold-standard biomarker for all metals (e.g., bone for lead). Levels are unlikely to vary significantly over time, given environmental persistence and stable diets in Bangladesh(20,55,71). Third, dietary intake and micronutrients were not measured, leaving potential for residual confounding, though the relatively homogeneous rural diet based on rice, fish, and vegetables may reduce variability(55,75). Fourth, both SES and metal concentrations were measured at baseline, raising concerns about temporality and the possibility that metals may act as both mediators and confounders; however, participants were aged ≥25 years and likely to have completed their SES attainment by enrollment. Fifth, selection bias could affect mediation estimates, but sensitivity analyses using IPCW and comparisons of complete-case and imputed data supported the robustness of our findings. Sixth, the study population was drawn from a highly arsenic-exposed region, which may limit generalizability. Sixth, we did not adjust for BMI or diabetes because these are intermediary variables in the SES–BP and metals–BP pathways. Finally, a key limitation of this study is the reliance on several assumptions, including no unmeasured confounding of SES-BP, SES-metal exposures, and metals-BP associations. This is unlikely to hold given the absence of factors such as air pollution, dietary sodium, and physical activity, which may bias the estimates. Nonetheless, sensitivity analyses suggest our mediation findings are robust to potential unmeasured confounding (not shown).

## 5. Conclusion

Our findings demonstrate socioeconomic differentials in blood pressure outcomes and inequalities in metal mixture exposures in Bangladesh. Higher education increases BP despite lowering metal exposures; the indirect effect mediated through metals was protective, but insufficient to counteract the overall harmful influence of higher education on BP. In contrast, the protective effect of SEP on BP outcomes was almost fully mediated by reduced metal exposures, suggesting that interventions targeting environmental mixtures, rather than changes in SEP itself, could yield substantial benefits. These results validate the mediating role of environmental mixtures in the SES–BP relationship and highlight the value of four-way decomposition analysis in uncovering pathways of environmental injustice and socioeconomic gradient in BP outcomes in LMICs.

## Data Availability

The data that has been used is confidential.

## Declaration of generative AI and AI-assisted technologies in the manuscript preparation process

During the preparation of this work, the authors used Grammarly in order to fix spelling and grammatical errors. After using this tool/service, the authors reviewed and edited the content as needed and take full responsibility for the content of the published article.

## Data Statement

Data is confidential.

## Supplementary Results

**Table S1:** Variables included in multiple imputation process (n=7000)

| Variables | Missing Values/Lost to Follow-up |  |
| --- | --- | --- |
|  | Count | Percent |
| **Covariates** |  |  |
| Sex | 0 | 0.0% |
| Age | 0 | 0.0% |
| Site | 0 | 0.0% |
| BMI | 0 | 0.0% |
| Medication for HBP | 0 | 0.0% |
| Treatment group | 0 | 0.0% |
| Smoking status | 0 | 0.0% |
| Betel quid use | 0 | 0.0% |
| Diabetic | 0 | 0.0% |
| **Socioeconomic Status** |  |  |
| Years of education | 0 | 0.0% |
| Ownership of agricultural land | 0 | 0.0% |
| Occupation | 0 | 0.0% |
| **Metal Concentration in Biomarkers** |  |  |
| Blood lead (µg/L) | 6014 | 85.9% |
| Blood selenium (µg/L) | 37 | 0.5% |
| Blood arsenic (µg/L) | 37 | 0.5% |
| Creatinine-adjusted Urinary arsenic (µg/g Cr) | 35 | 0.4% |
| **Blood Pressure Outcomes: Lost to Follow-up at 3<sup>rd</sup> Visit** |  |  |
| SBP at baseline | 0 | 0.0% |
| DBP at baseline | 0 | 0.0% |
| PP at baseline | 0 | 0.0% |
| MAP at baseline | 0 | 0.0% |
| SBP at follow-up 3 | 1077 | 15.4% |
| DBP at follow-up 3 | 1077 | 15.4% |
| PP at follow-up 3 | 1077 | 15.4% |
| MAP at follow-up 3 | 1077 | 15.4% |

**Table S2:** Comparing baseline metal mixtures exposure between complete and imputed data (N=5923)

|  | N | Missing (%) | Geometric Mean (95% CI) |  |  | Percentile |  |  |  |  |
| --- | --- | --- | --- | --- | --- | --- | --- | --- | --- | --- |
|  |  |  | Gmean | 2.5 % | 97.5% | Min | 25<sup>th</sup> | Med<br>ian | 75<sup>th</sup> | Max |
| Heavy Metals Concentration in Biomarkers |  |  |  |  |  |  |  |  |  |  |
| *Complete Cases Measured at Baseline* |  |  |  |  |  |  |  |  |  |  |
| Blood lead (µg/L) | 900 | 5023 (84.8%) | 76.9 | 73.1 | 80.9 | 2.0 | 42.0 | 88.0 | 141.0 | 354.0 |
| Blood selenium (µg/L) | 5894 | 29 (0.5%) | 130.4 | 129.8 | 131.1 | 64.0 | 115.0 | 130.0 | 147.0 | 927.0 |
| **Blood arsenic (µg/L)** | 5894 | 29 (0.5%) | 12.1 | 11.8 | 12.5 |  | 4.7 | 14.8 | 31.8 |  |
| **Creatinine-adjusted Urinary (µg/g Cr)** | 5902 | 21 (0.4%) | 379.7 | 368.2 | 391.7 | 11.3 | 141.4 | 468.5 | 1047.0 | 4816.3 |
| *Imputed Cases Measured at Baseline [pooled across 10 imputed datasets] \** |  |  |  |  |  |  |  |  |  |  |
| **Blood lead (µg/L)** | 5923 | Imputed | 67.4 | 66.0 | 68.8 | 2.0 | 35.5 | 70.3 | 128.0 | 354.0 |
| **Blood selenium (µg/L)** | 5923 | Imputed | 130.4 | 129.8 | 131.1 | 64.0 | 115.0 | 130.0 | 147.0 | 927.0 |
| **Blood arsenic (µg/L)** | 5923 | Imputed | 12.1 | 11.8 | 12.5 | 70.0 | 4.7 | 14.8 | 31.8 | 31.8 |
| **Creatinine-adjusted Urinary (µg/g Cr)** | 5923 | Imputed | 379.5 | 367.9 | 391.4 | 11.3 | 141.3 | 468.3 | 1046.5 | 4816.3 |
| *. |  |  |  |  |  |  |  |  |  |  |

**Table S3:**
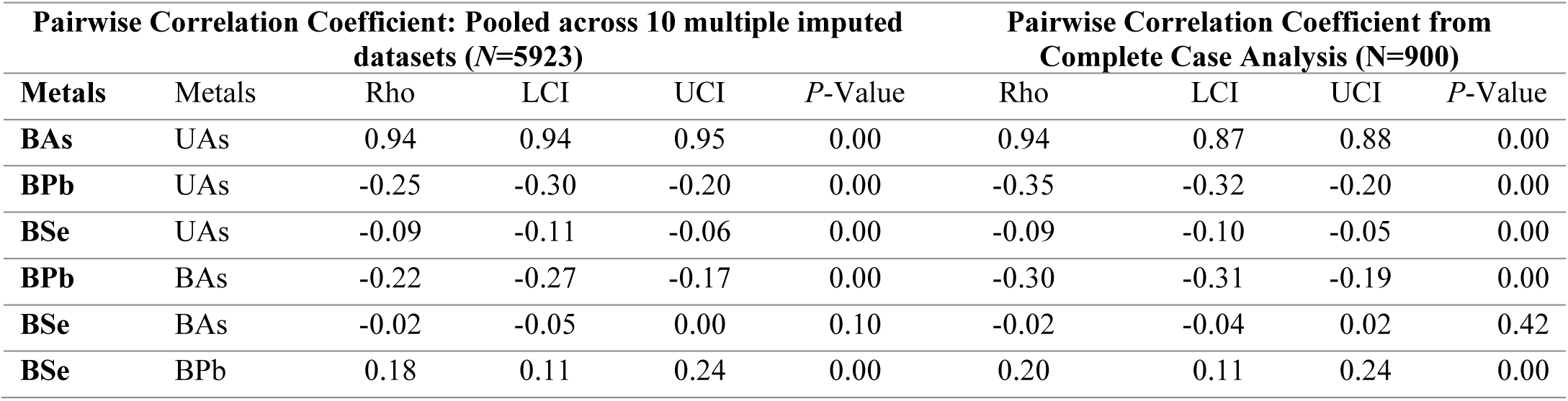
Comparison of Pairwise Spearman correlations among metal mixtures.

**Table S4:**
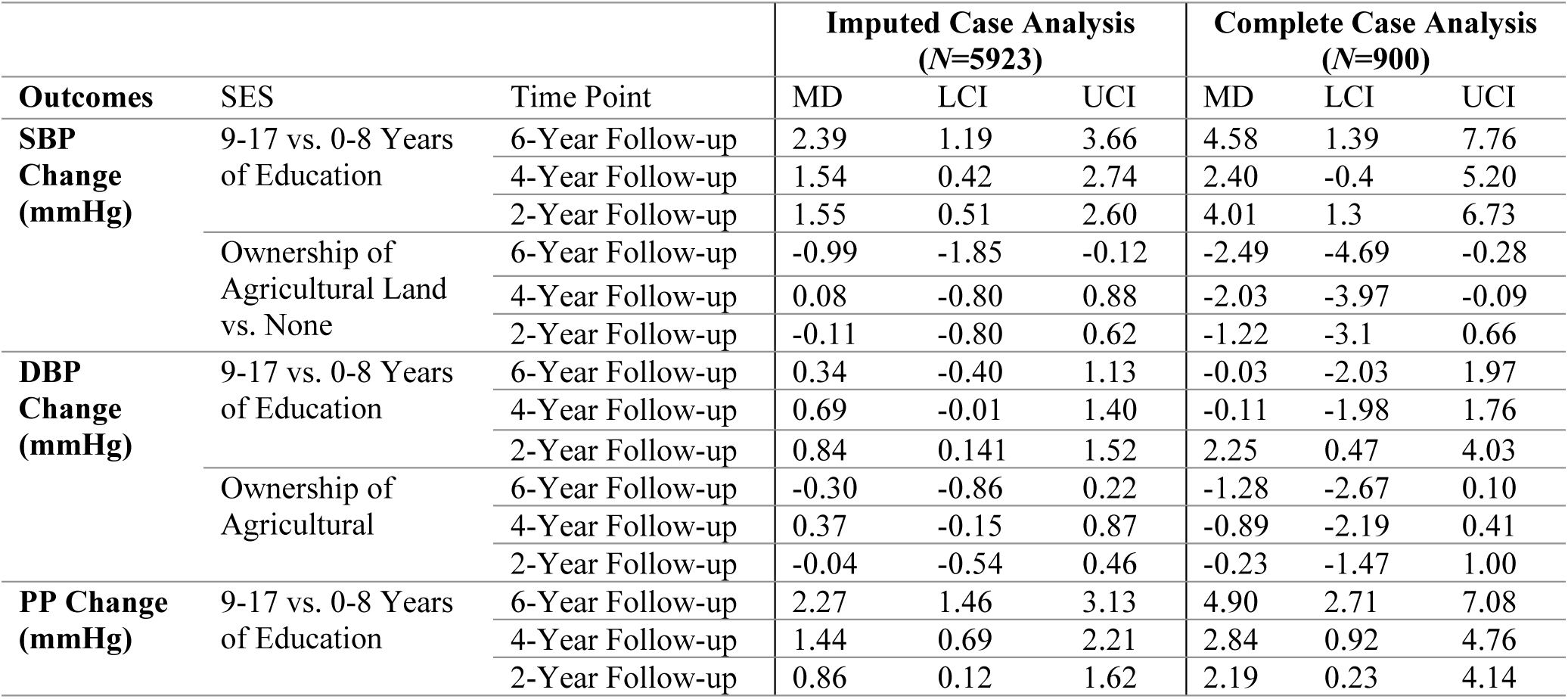
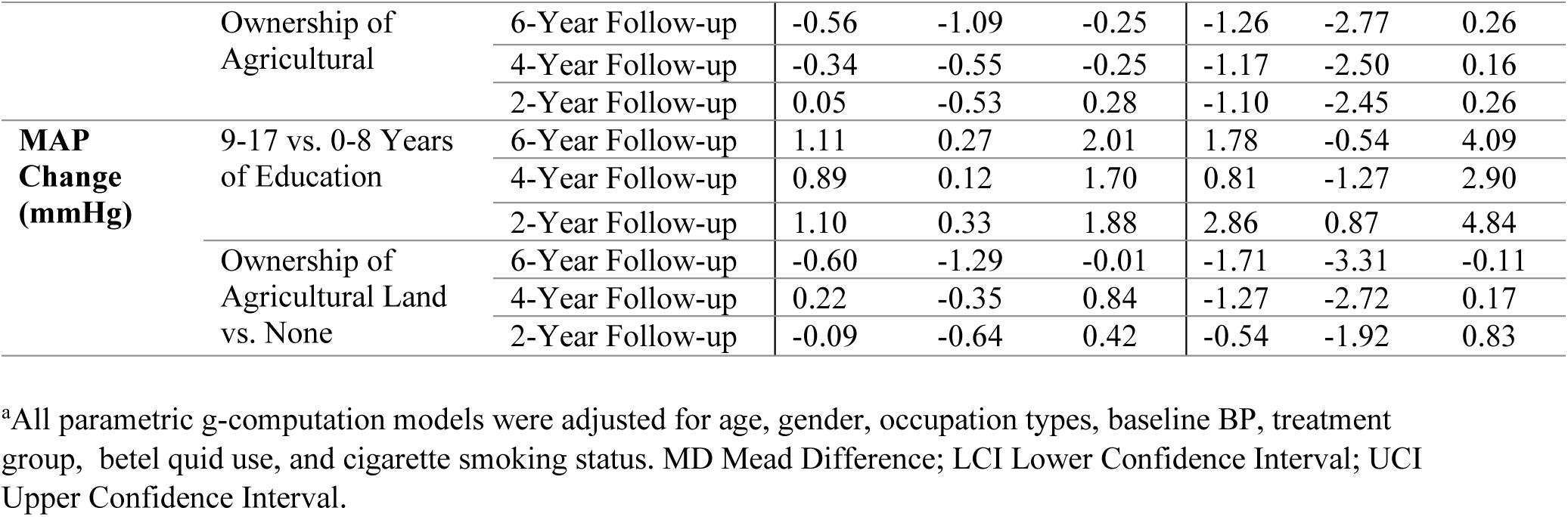
Marginal total effects of baseline SES indicators on BP Outcomes over six years of follow-up^a^.

**Table S5:**
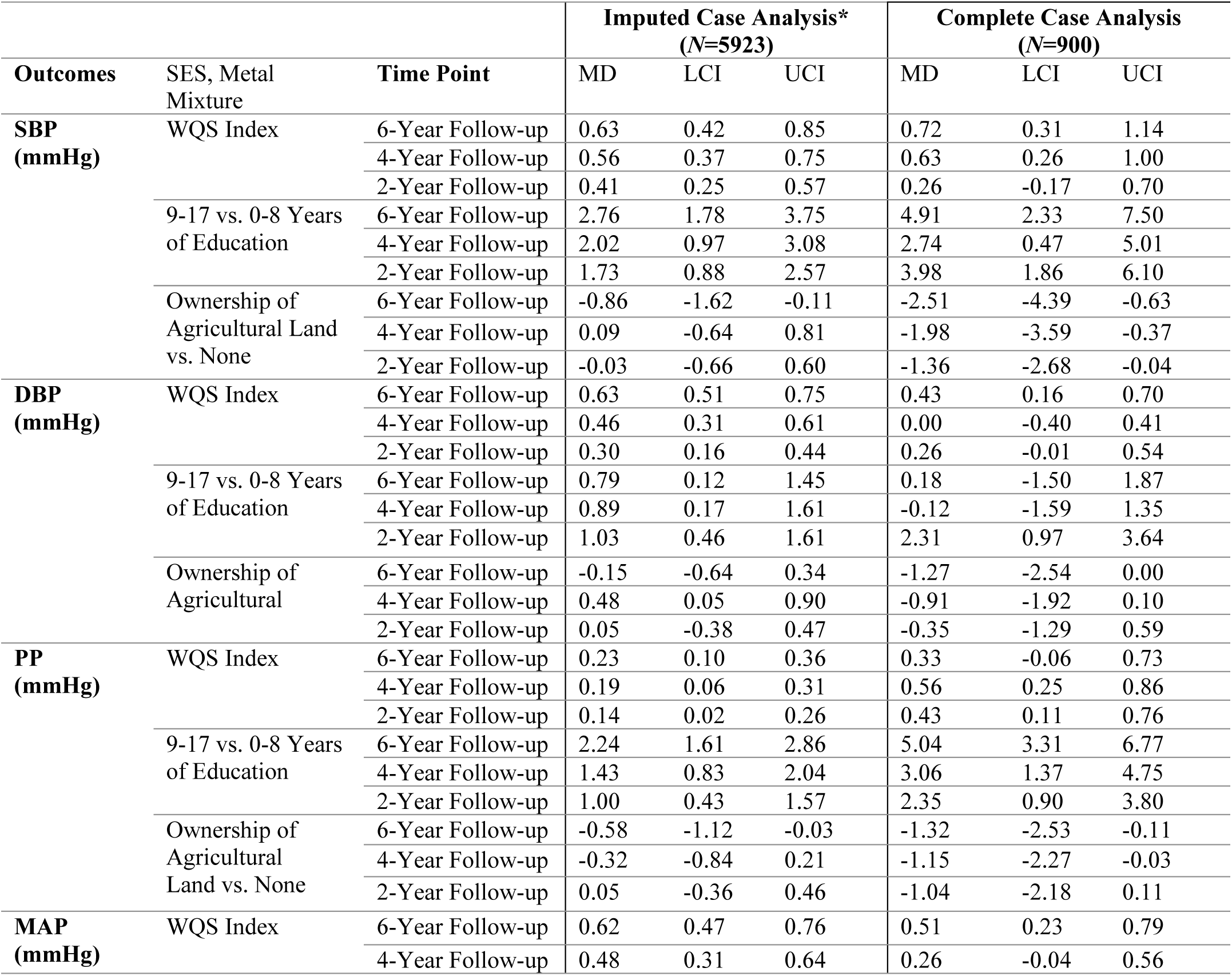
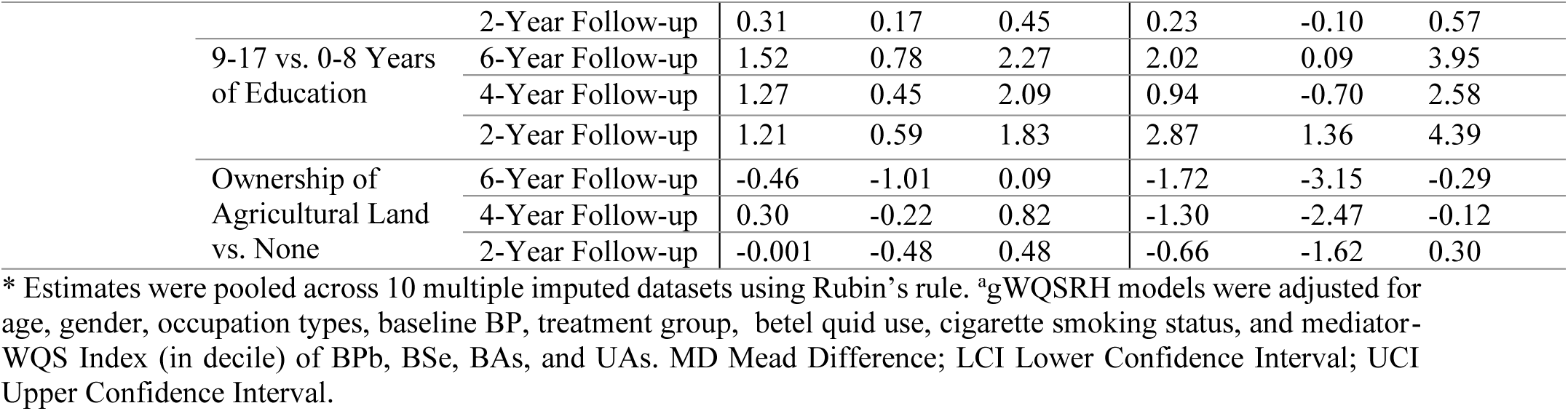
Direct effects of baseline SES and metal mixture exposure on BP Outcomes over six years of follow-up^a^.

**Table S6:** Marginal total effects of SES indicators on WQS metal mixture Index (in decile)

| SES Indicators | Imputed Case Analysis (N=5923) |  | Complete Case Analysis (N=900) |  |
| --- | --- | --- | --- | --- |
|  | Metal Mixture* (95% CI ) | Mean Difference (95% CI ) | Metal Mixture (95% CI ) | Mean Difference (95% CI ) |
| Educational Years and WQS metal mixture Index |  |  |  |  |
| 0-8 Years of Education | 4.47 (4.41, 4.52) | Reference | 4.47 (4.32, 4.63) | Reference |
| 9-11 Years of Education | 3.83 (3.67, 4.00) | -0.63 (-0.78, -0.48) | 3.62 (3.23, 4.04) | -0.86 (-1.21, -0.46) |
| Agricultural Land Ownership (ALO) and WQS metal mixture Index |  |  |  |  |
| Non-ALO | 4.47 (4.41, 4.52) | Reference | 4.47 (4.32, 4.63) | Reference |
| ALO | 4.17 (4.05, 4.29) | -0.30 (-0.40, -0.19) | 4.09 (3.79, 4.42) | -0.38 (-0.64, -0.12) |

CI: Confidence Interval. \*In decile of WQS metal mixture Index

**Table S7a:**
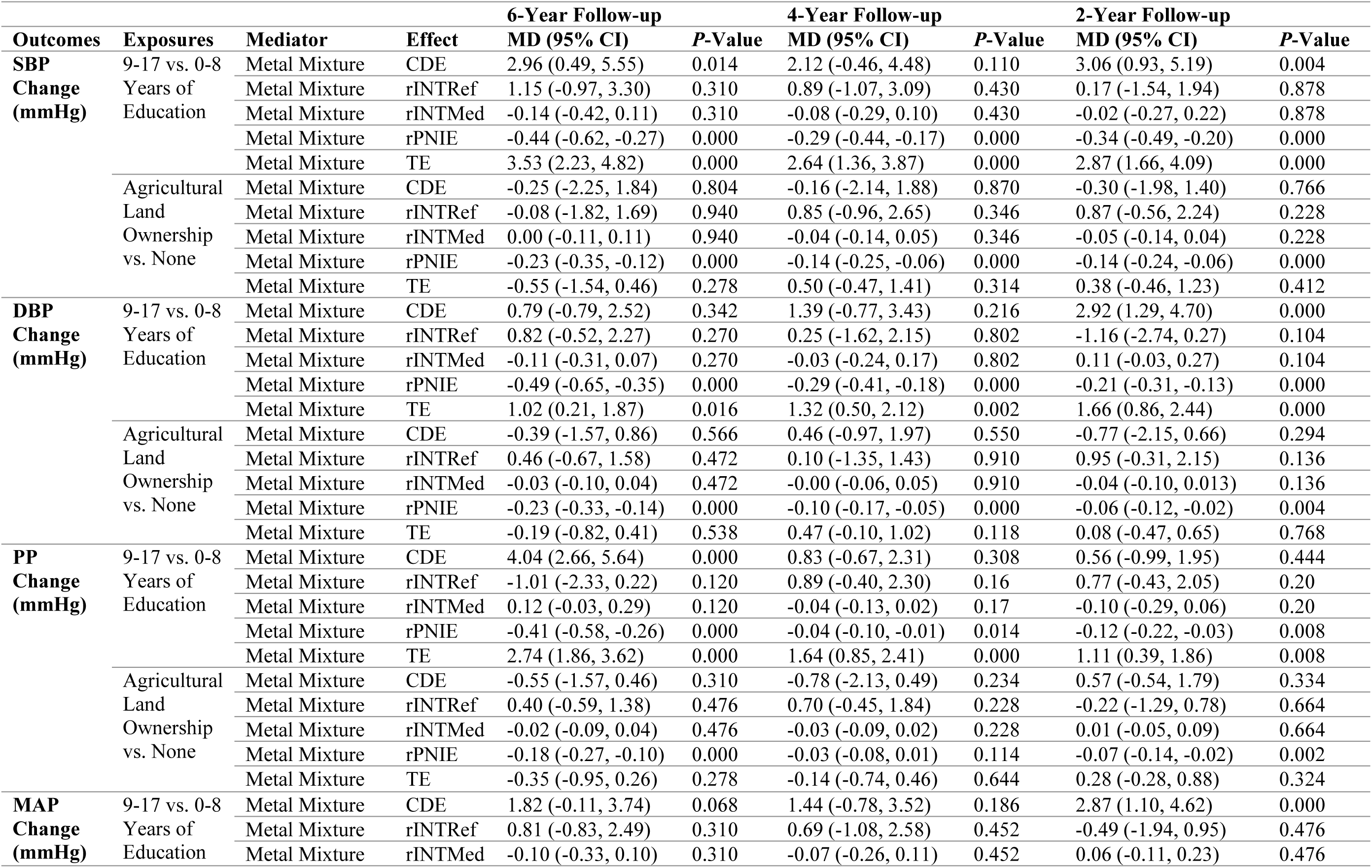
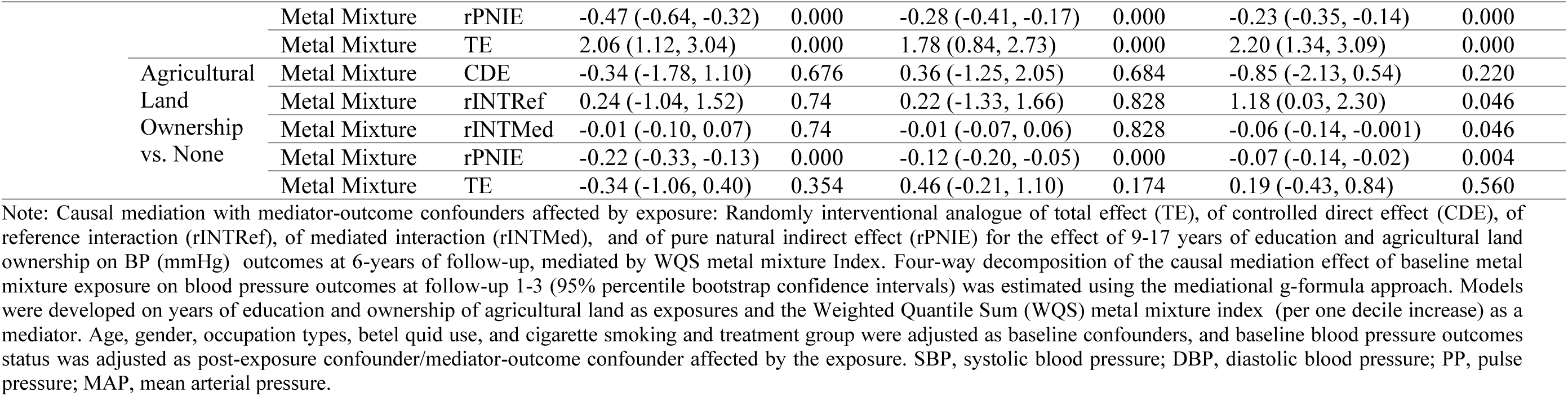
Randomly interventional analog of TE, of CDE, of rINTRef, of rINTMed, and of rPNIE for the effect of education and ALO on BP outcomes over 6 years of follow-up, mediated by WQS metal mixture Index (*N*=5923).

**Table S7b:**
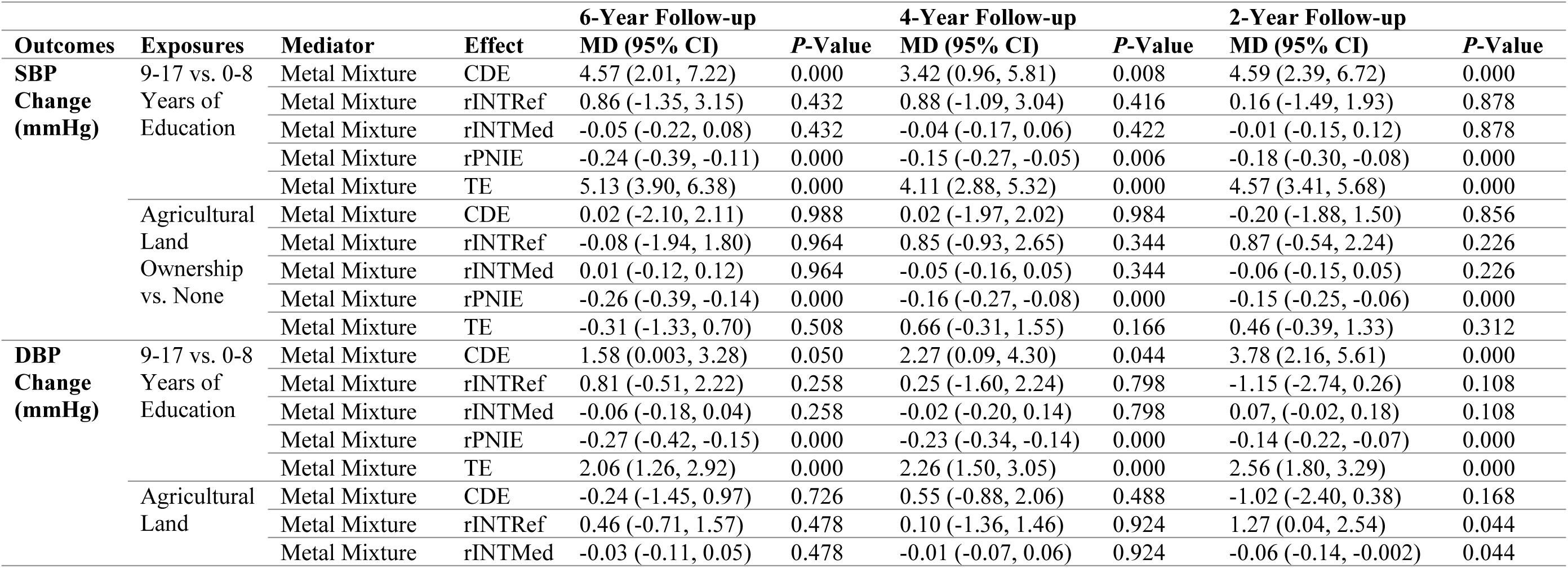
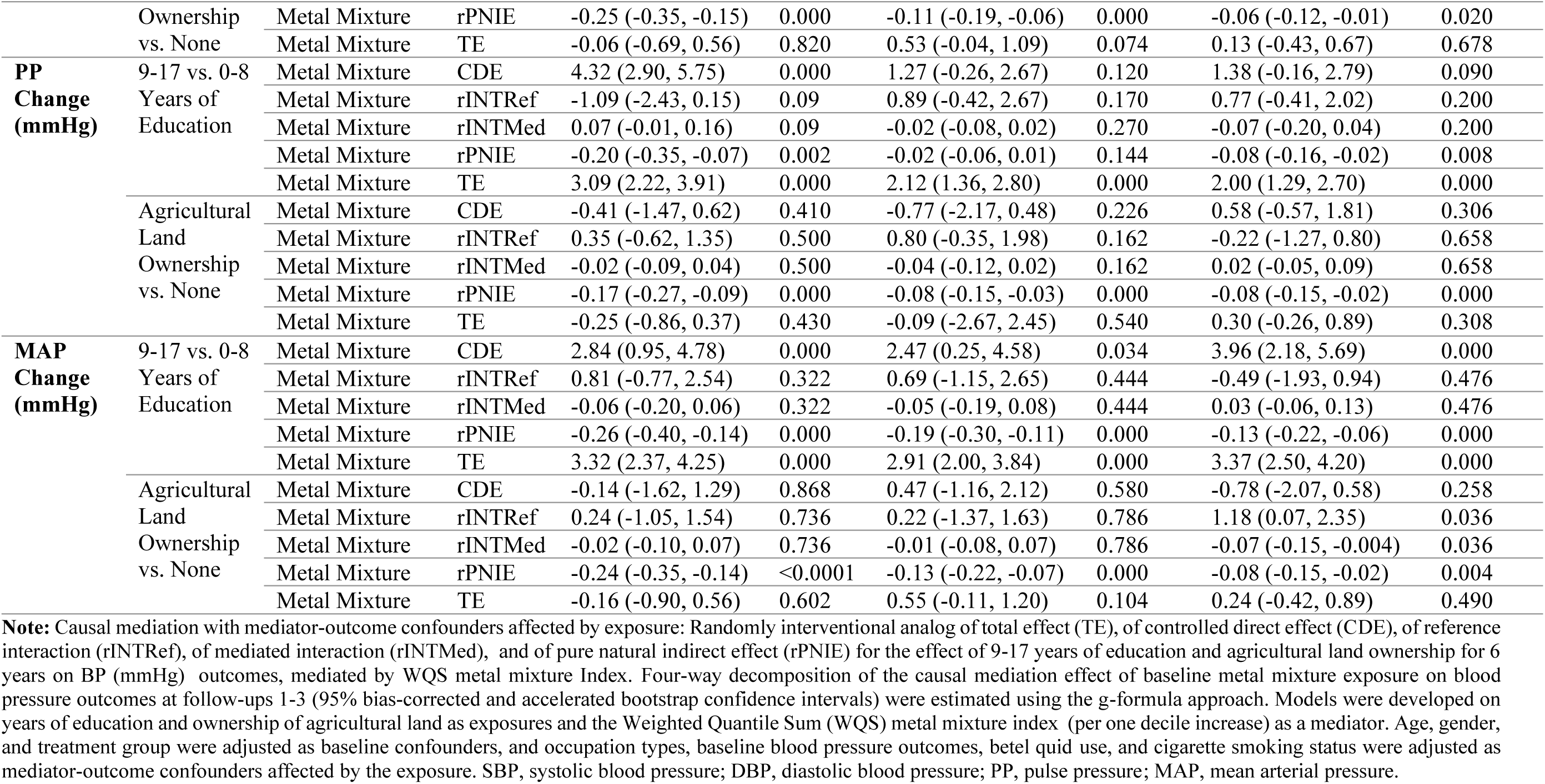
Randomly interventional analogue of TE, of CDE, of rINTRef, of rINTMed, and of rPNIE for the effect of education and ALO on BP outcomes over 6 years of follow-up, mediated by WQS metal mixture Index (*N*=5923).

**Table S8a:**
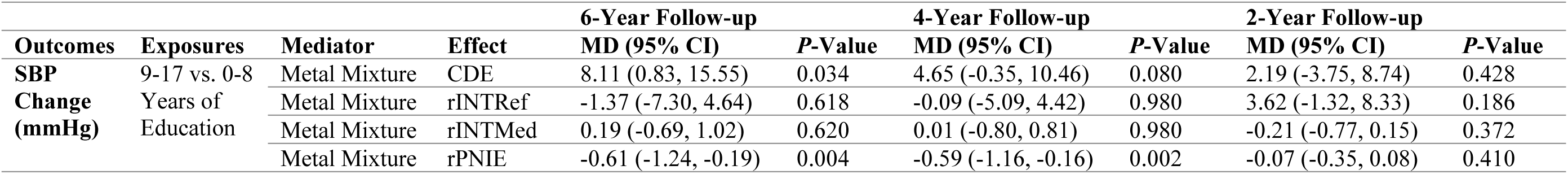
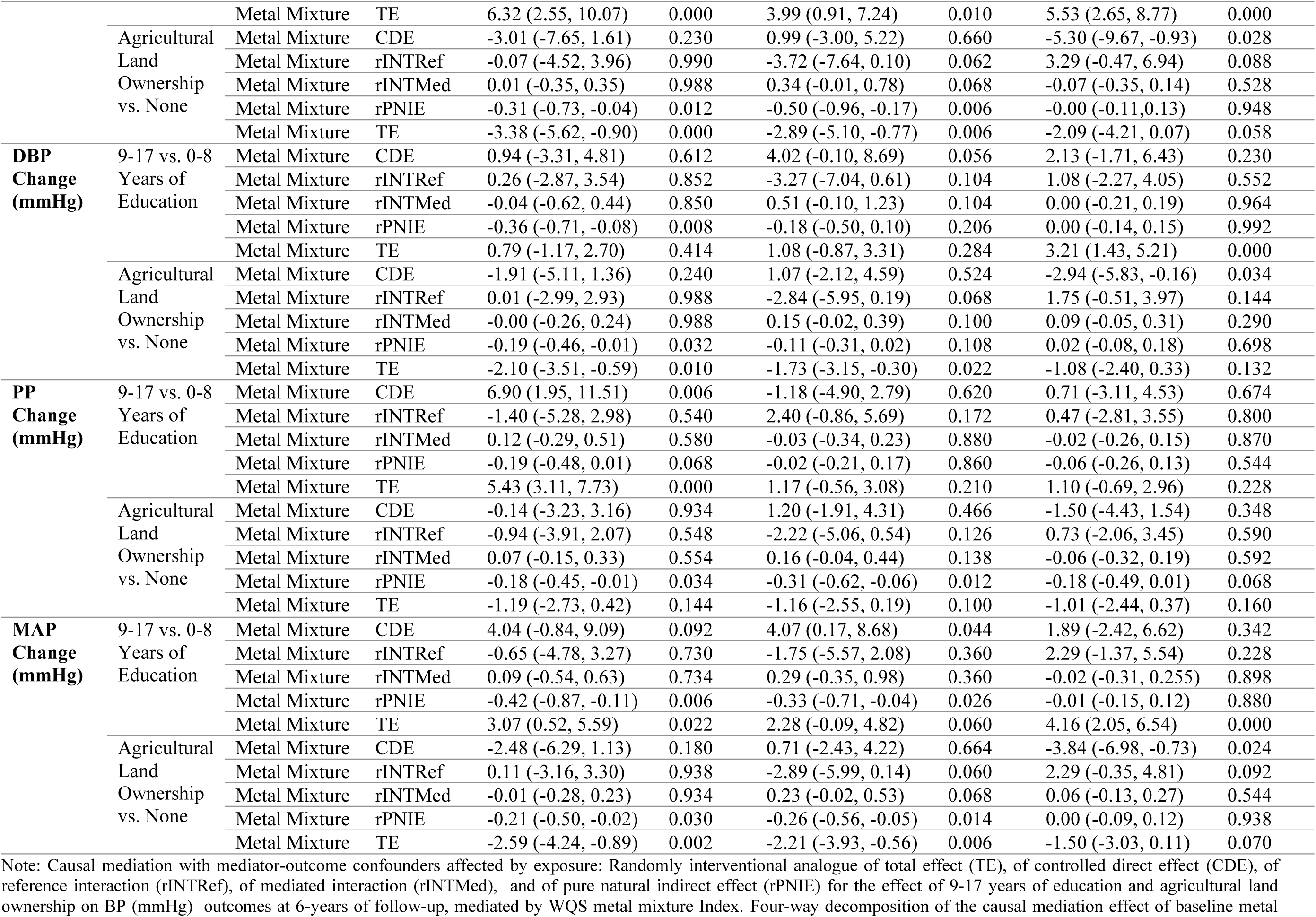
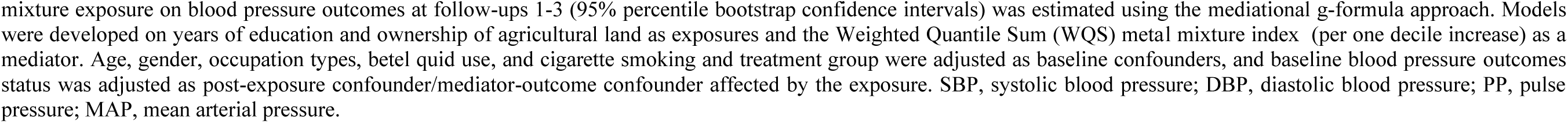
Randomly interventional analog of TE, of CDE, of rINTRef, of rINTMed, and of rPNIE for the effect of education and ALO on BP outcomes over 6 years of follow-up, mediated by WQS metal mixture Index (*N*=900)

**Table S8b:**
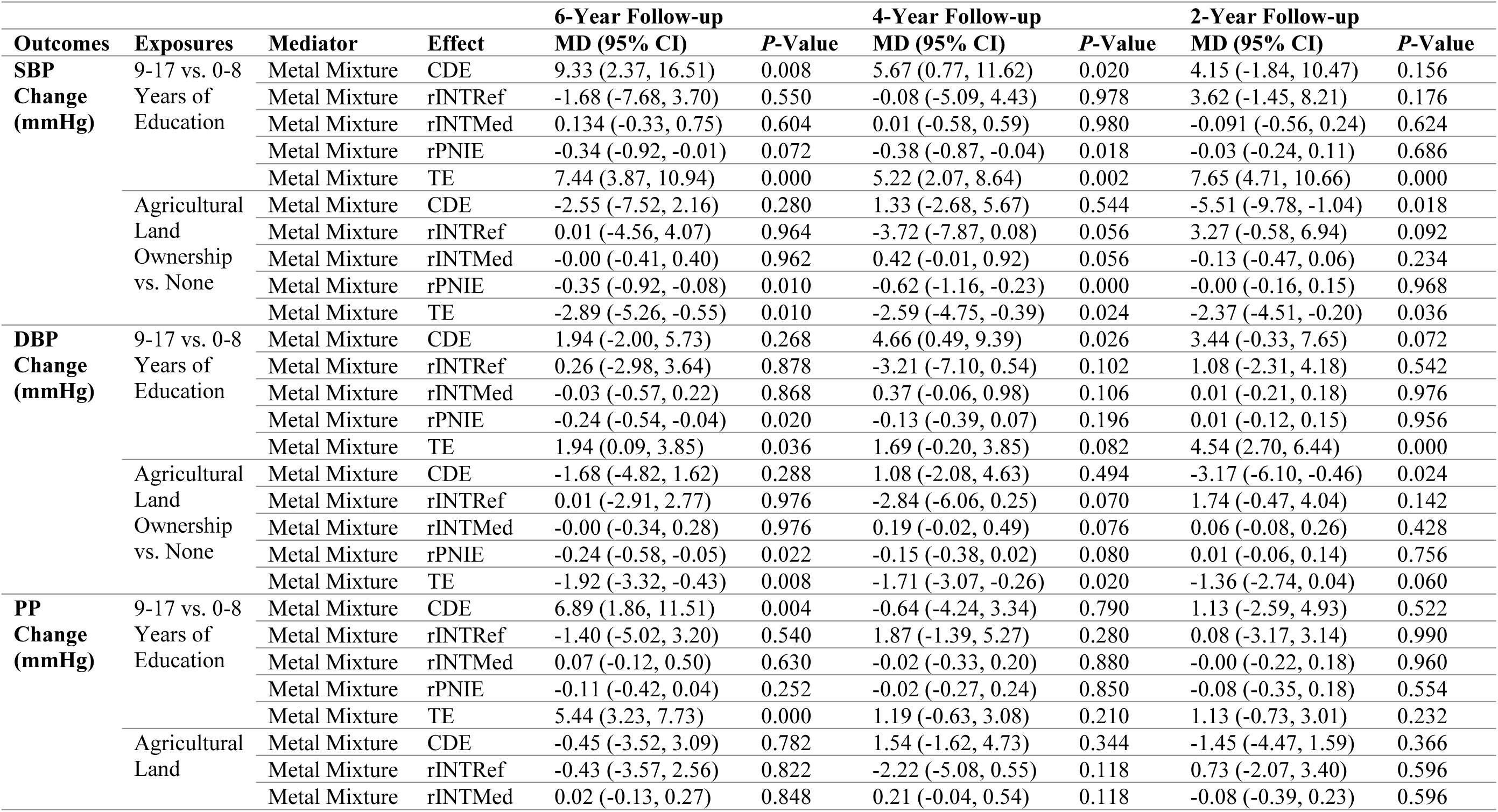
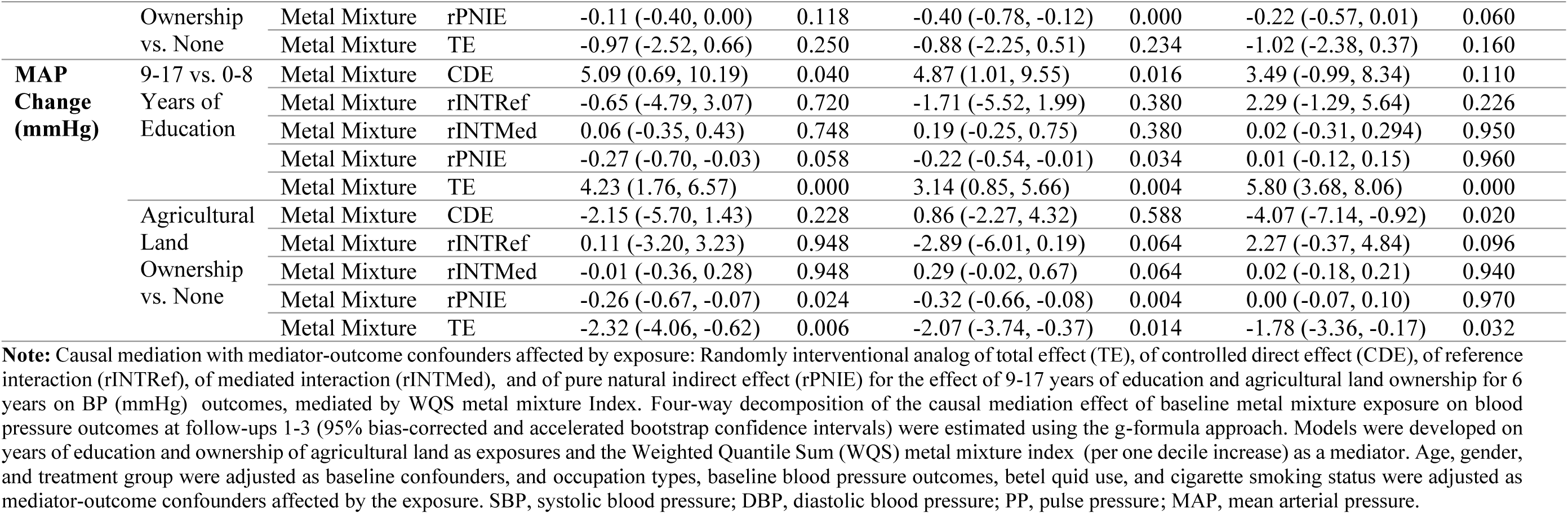
Randomly interventional analog of TE, of CDE, of rINTRef, of rINTMed, and of rPNIE for the effect of education and ALO on BP outcomes over 6 years of follow-up, mediated by WQS metal mixture Index (*N*=900)

**Table S9a:**
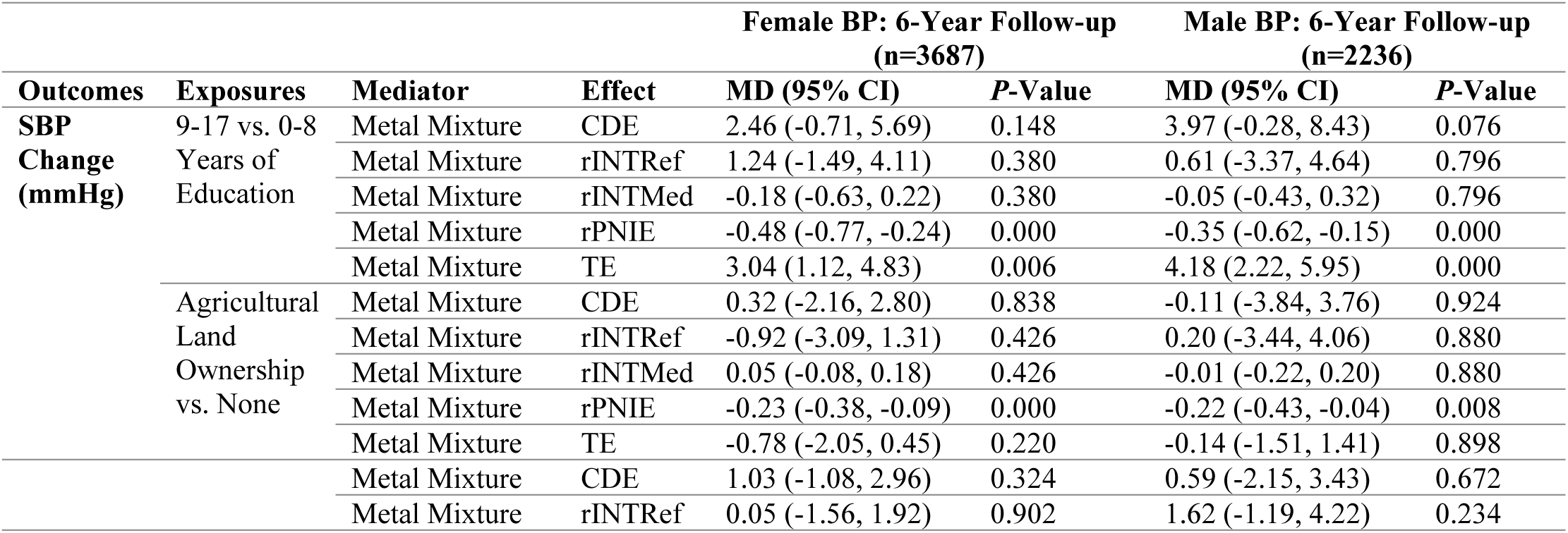
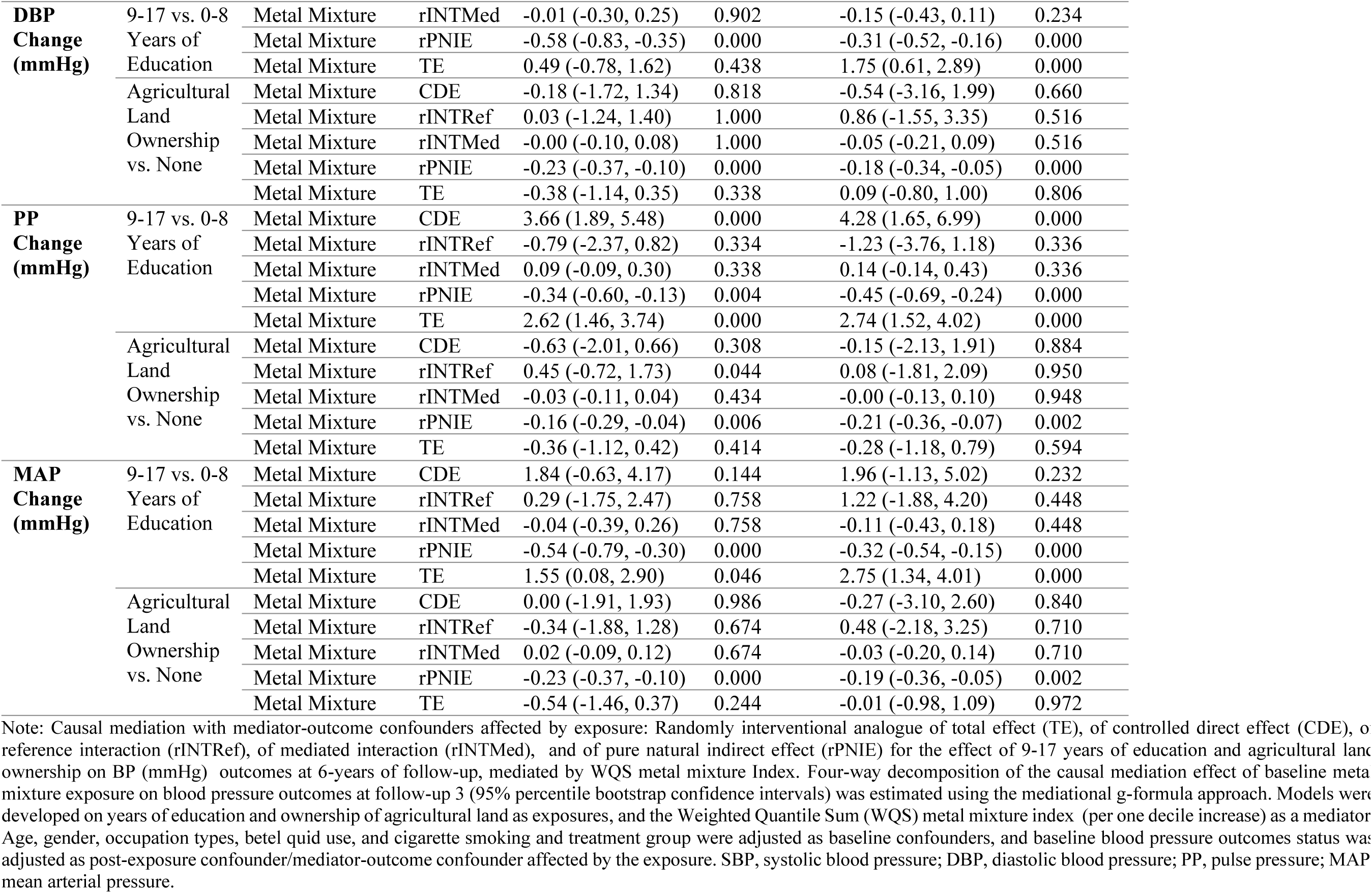
Gender-specific randomly interventional analog of TE, of CDE, of rINTRef, of rINTMed, and of rPNIE for the effect of education and ALO on BP outcomes over 6 years of follow-up, mediated by WQS metal mixture Index (*N*=5923).

**Table S9b:** Gender-specific randomly interventional analog of TE, of CDE, of rINTRef, of rINTMed, and of rPNIE for the effect of education and ALO on BP outcomes over 6 years of follow-up, mediated by WQS metal mixture Index (*N*=5923).

| Outcomes | Exposures | Mediator | Effect | Female BP: 6-Year Follow-up<br>(n=3687) |  | Male BP: 6-Year Follow-up<br>(n=2236) |  |
| --- | --- | --- | --- | --- | --- | --- | --- |
|  |  |  |  | MD (95% CI) | P-Value | MD (95% CI) | P-Value |
| **SBP**<br>**Change**<br>**(mmHg)** | 9-17 vs. 0-8 | Metal Mixture | CDE | 3.97 (0.74, 7.21) | 0.014 | 5.89 (1.63, 10.53) | 0.008 |
|  | Years of | Metal Mixture | rINTRef | 0.73 (-2.20, 3.73) | 0.608 | 0.55 (-3.47, 4.57) | 0.844 |
|  | Education | Metal Mixture | rINTMed | -0.06 (-0.34, 0.19) | 0.604 | -0.03 (-0.27, 0.18) | 0.842 |
|  |  | Metal Mixture | rPNIE | -0.28 (-0.54, -0.08) | 0.004 | -0.19 (-0.39, -0.03) | 0.006 |
|  |  | Metal Mixture | TE | 4.37 (2.51, 6.21) | 0.000 | 6.22 (4.44, 8.14) | 0.000 |
|  | Agricultural<br>Land | Metal Mixture | CDE | 0.47 (-2.08, 2.95) | 0.768 | 0.53 (-3.27, 4.48) | 0.822 |
|  |  | Metal Mixture | rINTRef | -0.80 (-3.06, 1.43) | 0.484 | -0.15 (-3.72, 3.84) | 0.966 |
|  | Ownership<br>vs. None | Metal Mixture | rINTMed | 0.05 (-0.10, 0.20) | 0.484 | 0.01 (-0.23, 0.26) | 0.966 |
|  |  | Metal Mixture | rPNIE | -0.23 (-0.43, -0.11) | 0.000 | -0.29 (-0.53, -0.08) | 0.002 |
|  |  | Metal Mixture | TE | -0.52 (-1.81, 0.63) | 0.372 | 0.09 (-1.22, 1.72) | 0.774 |
| **DBP**<br>**Change**<br>**(mmHg)** | 9-17 vs. 0-8 | Metal Mixture | CDE | 1.64 (-0.32, 3.46) | 0.108 | 1.76 (-0.97, 4.60) | 0.196 |
|  | Years of | Metal Mixture | rINTRef | 0.05 (-1.56, 1.97) | 0.900 | 1.58 (-1.22, 4.21) | 0.238 |
|  | Education | Metal Mixture | rINTMed | -0.005 (-0.19, 0.16) | 0.900 | -0.09 (-0.27, 0.07) | 0.242 |
|  |  | Metal Mixture | rPNIE | -0.33 (-0.60, -0.13) | 0.000 | -0.18 (-0.33, -0.05) | 0.004 |
|  |  | Metal Mixture | TE | 1.36 (0.15, 2.46) | 0.032 | 3.07 (2.02, 4.20) | 0.000 |
|  | Agricultural<br>Land | Metal Mixture | CDE | -0.03 (-1.65, 1.48) | 0.976 | -0.33 (-2.84, 2.19) | 0.814 |
|  |  | Metal Mixture | rINTRef | 0.03 (-1.27, 1.42) | 0.978 | 0.85 (-1.59, 3.39) | 0.504 |
|  | Ownership<br>vs. None | Metal Mixture | rINTMed | -0.002 (-0.10, 0.09) | 0.978 | -0.06 (-0.23, 0.10) | 0.504 |
|  |  | Metal Mixture | rPNIE | -0.23 (-0.39, -0.11) | 0.000 | -0.22 (-0.38, -0.06) | 0.000 |
|  |  | Metal Mixture | TE | -0.23 (-1.02, 0.49) | 0.498 | 0.24 (-0.62, 1.20) | 0.482 |
| **PP**<br>**Change**<br>**(mmHg)** | 9-17 vs. 0-8 | Metal Mixture | CDE | 3.91 (2.17, 5.65) | 0.000 | 4.57 (2.08, 7.34) | 0.000 |
|  | Years of | Metal Mixture | rINTRef | -0.81 (-2.32, 0.74) | 0.312 | -1.25 (-3.79, 1.14) | 0.31 |
|  | Education | Metal Mixture | rINTMed | 0.03 (-0.06, 0.17) | 0.420 | 0.08 (-0.07, 0.27) | 0.318 |
|  |  | Metal Mixture | rPNIE | -0.13 (-0.36, 0.07) | 0.164 | -0.26 (-0.46, -0.07) | 0.010 |
|  |  | Metal Mixture | TE | 3.01 (1.86, 4.14) | 0.000 | 3.14 (1.93, 4.39) | 0.000 |
|  | Agricultural<br>Land | Metal Mixture | CDE | -0.55 (-1.90, 0.69) | 0.352 | 0.04 (-1.98, 2.10) | 0.930 |
|  |  | Metal Mixture | rINTRef | 0.41 (-0.81, 1.60) | 0.482 | 0.01 (-1.96, 2.01) | 0.984 |
|  | Ownership<br>vs. None | Metal Mixture | rINTMed | -0.02 (-0.11, 0.05) | 0.486 | -0.00 (-0.12, 0.11) | 0.988 |
|  |  | Metal Mixture | rPNIE | -0.13 (-0.28, -0.03) | 0.004 | -0.23 (-0.37, -0.06) | 0.008 |
|  |  | Metal Mixture | TE | -0.29 (-1.10, 0.48) | 0.468 | -0.17 (-1.06, 0.88) | 0.810 |
|  |  | Metal Mixture | CDE | 2.63 (0.30, 4.76) | 0.030 | 3.43 (0.26, 6.66) | 0.032 |
|  |  | Metal Mixture | rINTRef | 0.28 (-1.79, 2.48) | 0.760 | 1.19 (-1.82, 4.16) | 0.430 |
| **MAP Change (mmHg)** | 9-17 vs. 0-8 Years of Education | Metal Mixture | rINTMed | -0.02 (-0.24, 0.16) | 0.760 | -0.07 (-0.26, 0.10) | 0.430 |
|  |  | Metal Mixture | rPNIE | -0.31 (-0.55, -0.12) | 0.000 | -0.19 (-0.35, -0.05) | 0.004 |
|  |  | Metal Mixture | TE | 2.58 (1.15, 3.86) | 0.000 | 4.37 (3.06, 5.71) | 0.000 |
|  | Agricultural Land Ownership vs. None | Metal Mixture | CDE | 0.19 (-1.76, 1.97) | 0.854 | -0.03 (-2.94, 2.86) | 0.976 |
|  |  | Metal Mixture | rINTRef | -0.33 (-1.91, 1.27) | 0.674 | 0.48 (-2.22, 3.28) | 0.702 |
|  |  | Metal Mixture | rINTMed | 0.02 (-0.09, 0.13) | 0.674 | -0.03 (-0.23, 0.15) | 0.702 |
|  |  | Metal Mixture | rPNIE | -0.22 (-0.40, -0.11) | 0.000 | -0.24 (-0.42, -0.06) | 0.000 |
|  |  | Metal Mixture | TE | -0.35 (-1.29, 0.50) | 0.406 | 0.18 (-0.77, 1.34) | 0.594 |

**Note:** Causal mediation with mediator-outcome confounders affected by exposure: Randomly interventional analog of total effect (TE), of controlled direct effect (CDE), of reference interaction (rINTRef), of mediated interaction (rINTMed), and of pure natural indirect effect (rPNIE) for the effect of 9-17 years of education and agricultural land ownership for 6 years on BP (mmHg) outcomes, mediated by WQS metal mixture Index. Four-way decomposition of the causal mediation effect of baseline metal mixture exposure on blood pressure outcomes at follow-up 3 (95% bias-corrected and accelerated bootstrap confidence intervals) were estimated using the g-formula approach. Models were developed on years of education and ownership of agricultural land as exposures and the Weighted Quantile Sum (WQS) metal mixture index (per one decile increase) as a mediator. Age, gender, and treatment group were adjusted as baseline confounders, and baseline blood pressure outcomes, occupation types, betel quid use, and cigarette smoking status were adjusted as mediator-outcome confounders affected by the exposure. SBP, systolic blood pressure; DBP, diastolic blood pressure; PP, pulse pressure; MAP, mean arterial pressure.

**Table S10a:**
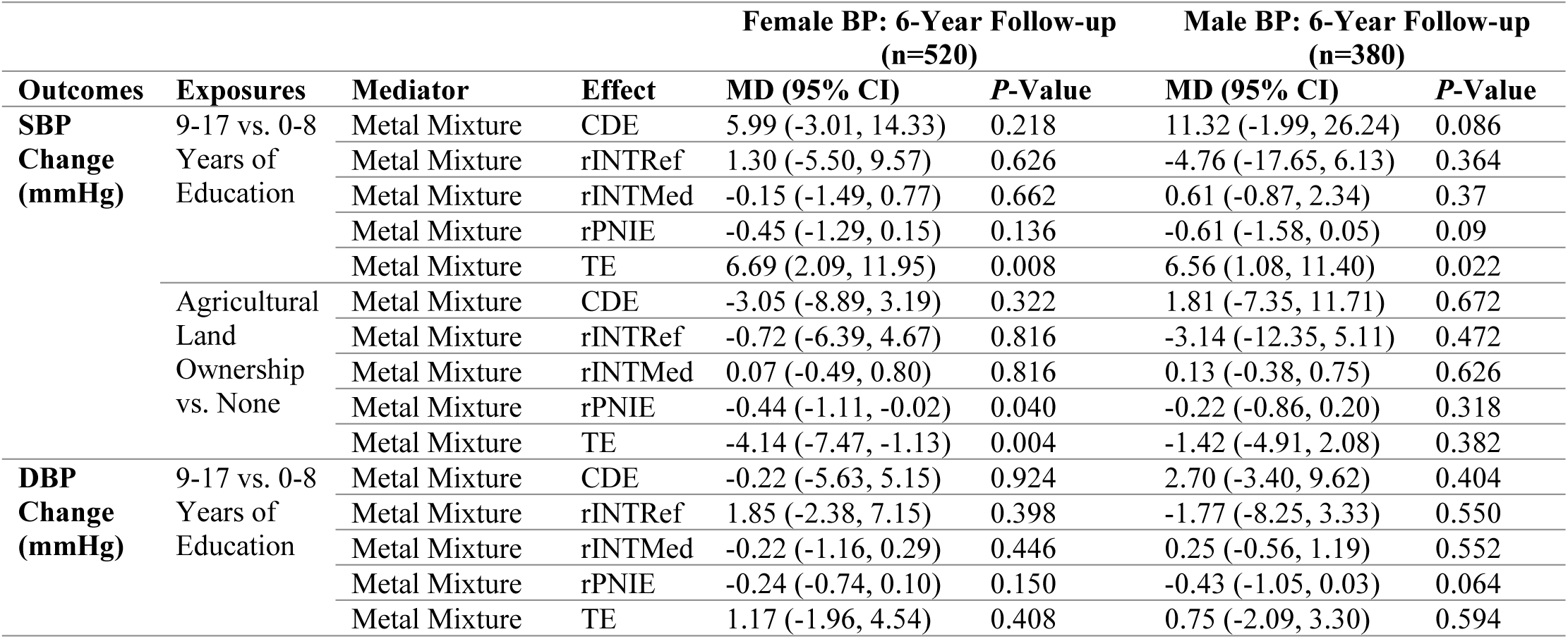
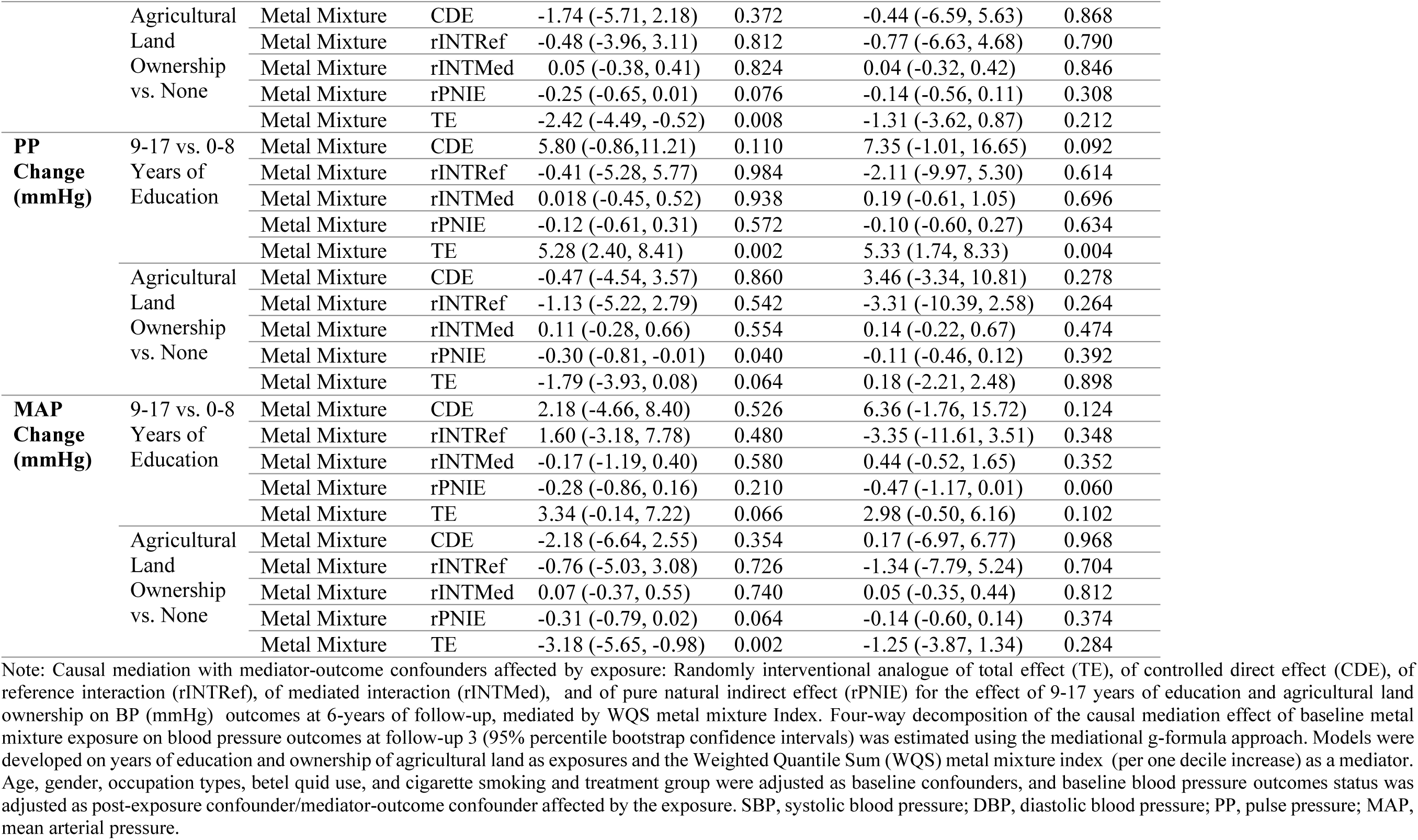
Gender-specific randomly interventional analog of TE, of CDE, of rINTRef, of rINTMed, and of rPNIE for the effect of education and ALO on BP outcomes over 6 years of follow-up, mediated by WQS metal mixture Index (*N*=900).

**Table S10b:**
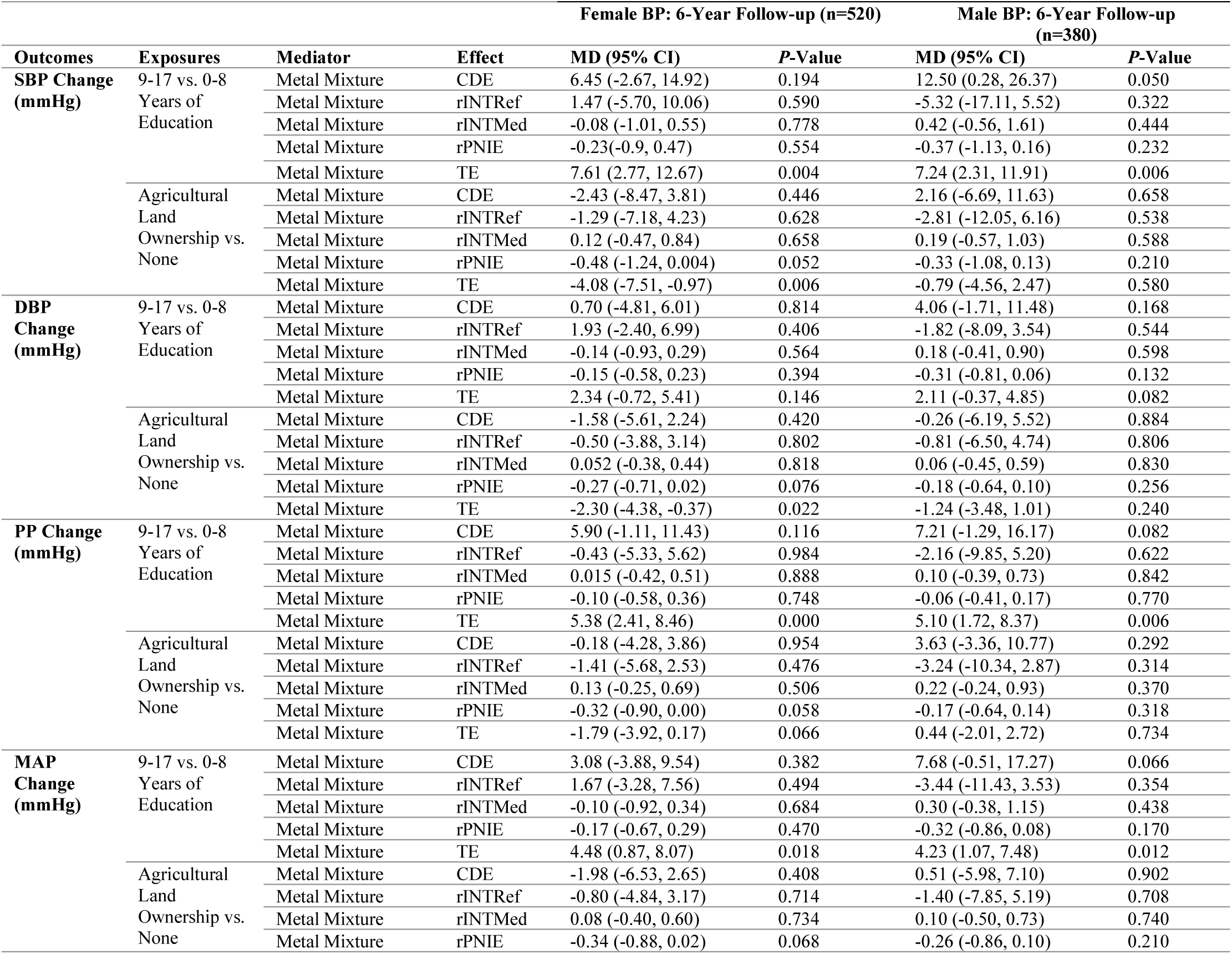
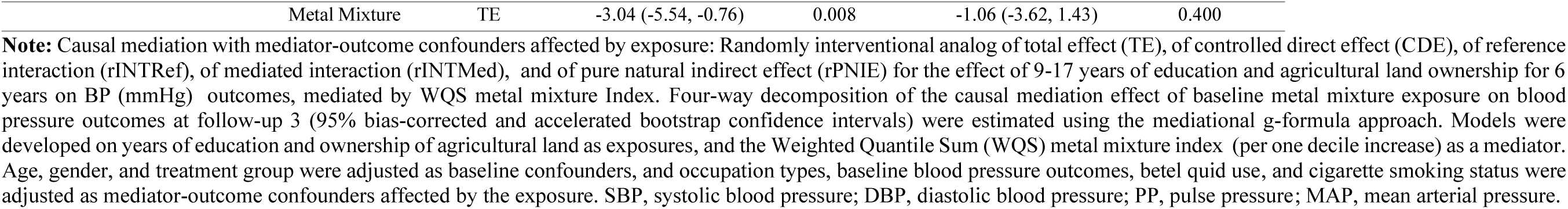
Gender-specific randomly interventional analog of TE, of CDE, of rINTRef, of rINTMed, and of rPNIE for the effect of education and ALO on BP outcomes over 6 years of follow-up, mediated by WQS metal mixture Index (*N*=900).

**Figure S1:**
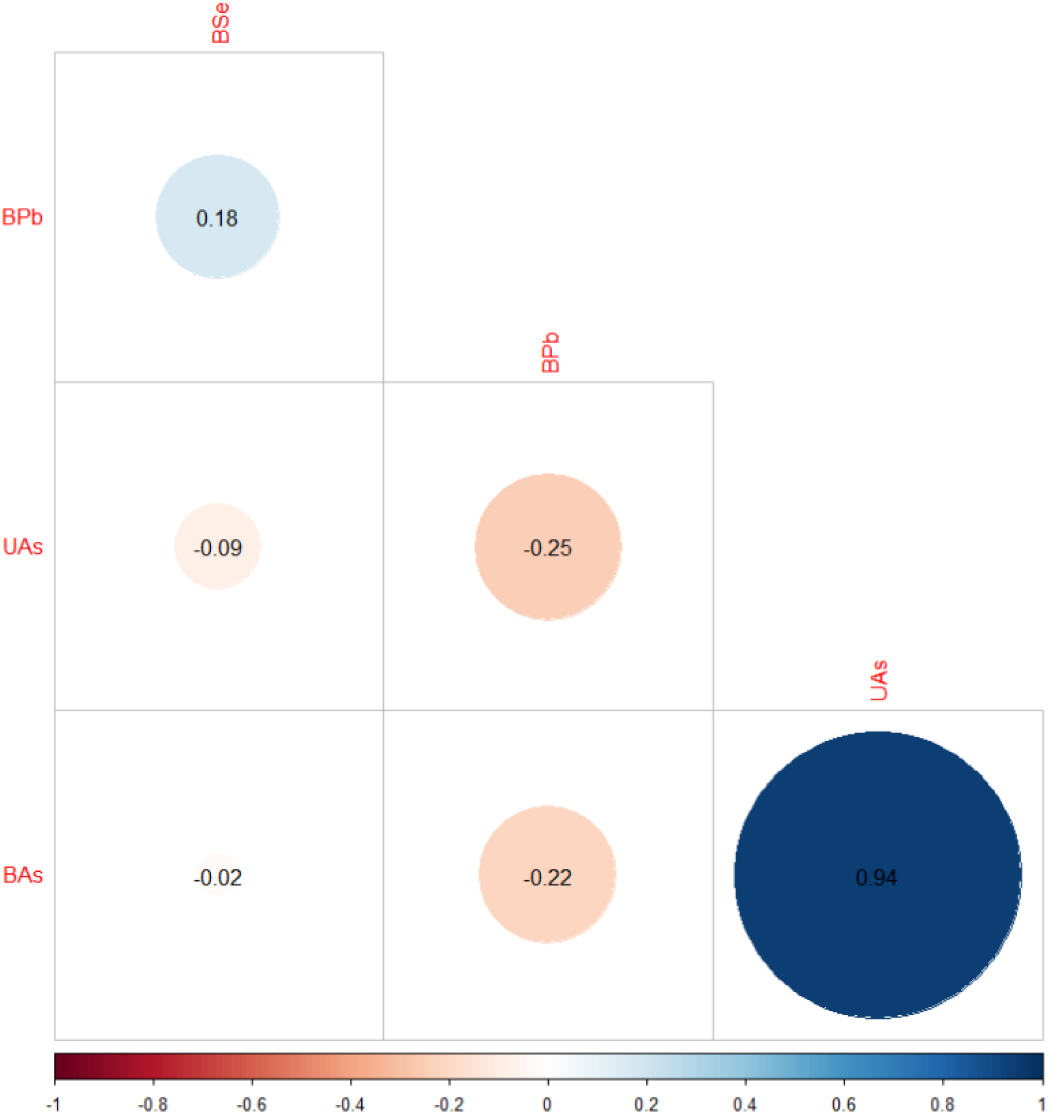
Pairwise Spearman correlations among metal mixtures (*N*=5923)

**Figure S2:**
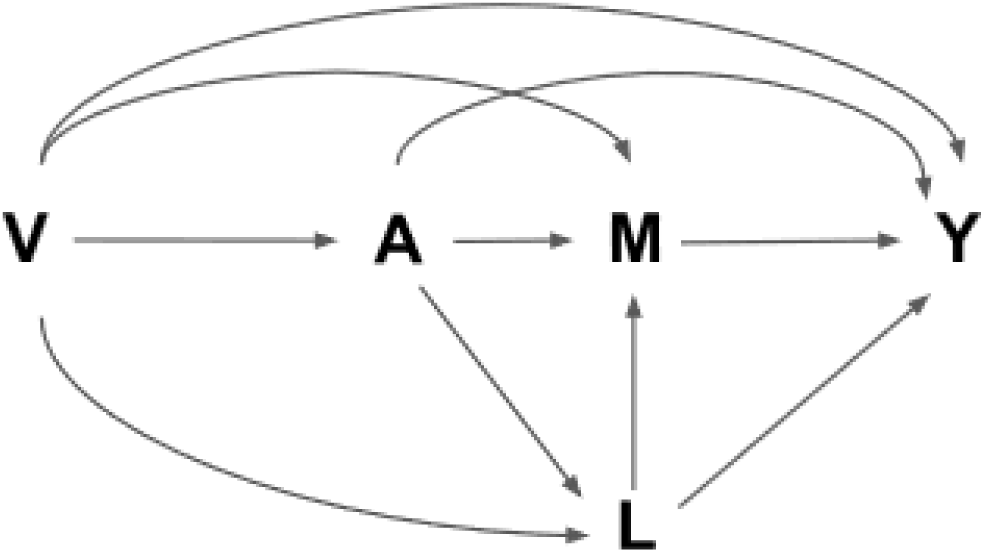
Causal Mediation Analysis with Mediator-Outcome Confounder L Affected by Exposure A **Note: A** (Exposures: Years of education and agricultural land ownership)**; Y** (Outcomes: SBP, DBP, PP, and MAP over 6 years of follow-up); **M** (Mediator: WQS metal mixture Index of BPb, BSe, BAs, and UAs); **V (**Baseline confounders not affected by exposure: age, gender, and treatment group); **L** (Mediator-outcome confounders affected by exposure: baseline blood pressure and/, betel quid use, smoking status, and occupational group).

